# Comparing Cadence vs. Machine Learning based Physical Activity Intensity Classifications: Variations in the Associations of Physical Activity with Mortality

**DOI:** 10.1101/2024.03.23.24304766

**Authors:** Le Wei, Matthew N. Ahmadi, Raaj Kishore Biswas, Stewart G. Trost, Emmanuel Stamatakis

## Abstract

**Background:** Step cadence-based and machine-learning (ML) methods have been used to classify physical activity (PA) intensity in health-related research. This study examined the association of intensity-specific PA daily duration with all-cause (ACM) and CVD mortality varied among cadence-based and ML methods in 68,561 UK Biobank participants.

**Methods:** The two-stage-ML method categorized activity type and then intensity. The one-level-cadence method (1LC) derived intensity duration using all detected steps and cadence thresholds of ≥100 steps/min (moderate intensity) and ≥130 steps/min (vigorous intensity). The two-level-cadence method (2LC) detected ambulatory activities and then steps with the same cadence thresholds.

**Results:** The 2LC exhibited the most pronounced association at the lower end of the duration, e.g., the 2LC showed the smallest minimum moderate-to-vigorous PA (MVPA) duration (amount associated with 50% of optimal risk reduction) (2LC vs 1LC vs ML, 2.8 minutes/day [95% CI: 2.6, 2.8] vs 11.1 [10.8, 11.4] vs 14.9 [14.6, 15.2]) while exhibiting similar corresponding ACM hazard ratio (HR) among methods (HR: 0.83 [95% CI: 0.78, 0.88] vs 0.80 [0.76, 0.85] vs 0.82 [0.76, 0.87]). The ML elicited the greatest mortality risk reduction, e.g., for VPA-ACM association, 2LC vs 1LC vs ML: median, 2.0 minutes/day [95% CI: 2.0, 2.0] vs 6.9 [6.9, 7.0] vs 3.2 [3.2, 3.2]; HR, 0.69 [0.61, 0.79] vs 0.68 [0.60, 0.77] vs 0.53 [0.44, 0.64]. After standardizing duration, the ML exhibited the most pronounced associations, e.g., for MPA-CVD mortality, 2LC vs 1LC vs ML, standardized minimum-duration: -0.77 vs - 0.85 vs -0.94; HR 0.82 [0.72, 0.92] vs 0.79 [0.69, 0.90] vs 0.77 [0.69, 0.85].

**Conclusion:** The 2LC exhibited the most pronounced association with mortality at the lower end of the duration. The ML method provided the most pronounced association with mortality after standardizing the durations.

## 1. Introduction

Current PA guidelines recommend weekly 150-300 moderate-to-vigorous intensity physical activity (MVPA) mins, or 75-150 vigorous intensity PA (VPA) mins and emphasize health benefits of all PA bouts regardless of their duration^1,2^. However, most of these guidelines were derived from self-reports which typically fail to recognize the health value of short PA bout (e.g., < 10-minute bout)^3^, thus affecting the accuracy of the estimates of the health impact of different PA intensities. Recent studies have shown that the short intermittent activity bursts were beneficially associated with long term health risks (e.g., all-cause^4,5^, CVD^4^ and cancer mortality risk^4^, and CVD^5^ and cancer^6^ incidence). The advancements in wearables measurement technology and methods for processing and analyzing large volumes of accelerometer data provide researchers with multiple options for classifying PA intensity within the short bout. Besides the more recent machine learning (ML) classification models ^7^, other methods include cut-points or thresholds based on proprietary activity counts ^8^ or gravitational units ^9^, or step cadence thresholds ^10^.

The step cadence of ≥ 100 steps/min has been examined as a heuristic threshold for moderate-to-vigorous intensity for adults in both treadmill and overground walking ^10–14^. Similarly, a cadence of ≥ 130 steps/min has been proposed as the heuristic threshold for vigorous intensity among young and middle-aged adults^10,11^. The step cadence methods are feasible with any accelerometer providing researchers with a relatively simple method to estimate the association of duration of PA across intensities with health outcomes in free-living samples. Prior studies have applied cadence thresholds to examine the association of PA intensities duration with various health outcomes ^15–21^. For instance, a harmonized meta-analysis including 15 international accelerometry cohorts (13 waist, 1 thigh, 1 wrist) found that the association of the duration of moderate or higher intensity (defined as ≥ 100 steps/min) with mortality was attenuated after adjusting for total step counts, albeit remained significant ^15^. Moreover, step is the fundamental unit of the ambulatory PA, step cadence-based thresholds may help people understand and monitor their required higher intensity PA minutes (e.g., at least 150 MPA minutes per week) ^1,2,22^ and support more people to be sufficiently active ^23^ .

ML methods have gained considerable attention in PA research recent years due to the increasing application of accelerometers and accessibility to raw data (gravitational acceleration) ^24^. By training a model with “features” of the accelerometer signal, such as time and frequency domain, that are extracted from the raw data, ML models classify patterns in the raw accelerometer signal corresponding to an activity type or intensity ^24^. ML algorithm has the advantage of high accuracy in identifying different PAs and intensities in both lab and free-living environments ^7,25,26^. Recent observational studies applying the ML method showed that approximately daily 2-3 short bouts or 3-4 minutes of vigorous intermittent lifestyle PA (VILPA) or 1-5 MV-ILPA minutes were associated with substantially reduced mortality risk, and cancer and CVD incidence^4–6^, amounts that are considerably lower than current major PA guidelines recommendations (at least 75 weekly VPA mins or 150 MPA mins) ^1,2,22^ primarily based on questionnaire-based data. Prior research indicated that the ML method demonstrated significantly higher agreement with ground-truth PA intensity than cut-point (counts) methods for preschool children in free-living environment ^27^.

A recent unpublished study of ours ^28^ indicated that the intensity-specific PA daily duration estimates based on step cadence thresholds were substantially different to those derived using the ML methods (mean absolute difference for MVPA: 24.2 mins/day). Such differences prompt the question of whether the PA intensity-mortality associations differ by intensity classification method in free-living conditions. To our knowledge, no previous study has examined the extent to which association between the duration of the PA intensity and mortality varies according to multiple methods derived by processing the accelerometer data to estimate PA intensity. Such information is important for guiding analytic and resource allocation decisions in observational research and for understanding the role of PA intensity classification in health research. The purpose of this study was to compare the association of intensity-specific daily PA duration with all-cause and CVD mortality, derived from two step cadence-based and one ML method, in a large UK cohort with wrist-worn accelerometer data.

## 2. Materials and Methods

### 2.1 Participants

The UK Biobank is a large prospective cohort with 502,616 aged 40-69 years UK adults recruited between 2006 and 2010 ^29^. Participants completed baseline measurements and provided written consent to use their data. The ethical approval was provided by the UK National Health Service, National Research Ethics Service (Ref 11/NW/0382).

From 2013 to 2015, 103,684 UK biobank participants were sent and wore the Axivity AX3 wrist-worn triaxial accelerometer (Newcastle upon Tyne, UK) on their dominant wrist for 7 days continuously. The AX3 was initialized to capture triaxial acceleration data at a sampling frequency of 100 Hz and a dynamic range of ±8 g. A monitoring day was considered valid if the wear time exceeded 16 hours. Participants returned the devices by mail and the data were calibrated ^30^ and non-wear periods were identified ^31^. Participants needed at least four valid monitoring days including a minimum of one valid weekend day to be included in our study. As previous study ^7^, we excluded participants with missing covariate data, CVD and cancer history (ascertained through hospital admission records), and outcome events occurring within one year after the PA assessment. In total, 68,561 participants were included in our study (**Supplementary figure 1**).

### 2.2 Physical activity intensity classification methods

#### 2.2.1 Machine learning method

This two-stage classifier first applied a validated random forest (RF) classifier trained on 48 time and frequency domain features to categorize each 10-second window into one of the four PA classes: sedentary (sitting still, reclined, etc.), standing utilitarian movements (standing stationary/active standing: washing dishes, ironing a shirt, etc.), walking activities (active commuting, ambulatory gardening, etc.), or running/energetic activity (chasing around with children, etc.) ^7^. Time windows classified as walking were assigned to different PA intensity bands using ambulatory acceleration thresholds and those classified as running/energetic activities were classified as vigorous (≥ 6 METs). Time windows classified as walking with normalized gravitational units < 100 milli g’s were considered LPA (<3 METs) and those classified as walking with normalized gravitation units ≥100 milli g and < 400 milli g were considered MPA (3-6 METs). The time windows classified as walking with normalized gravitation units ≥ 400 milli g walking activities were considered vigorous PA (VPA) (≥6 METs) ^32,33^. Windows classified as sedentary behaviors were considered sedentary (< 1.5 METs). We calculated the mean daily duration across intensities by averaging over valid monitoring days.

#### 2.2.2 One-level cadence method

We classified PA intensities using a validated one-level cadence method ^10^ (1LC). We implemented an open-source Windowed Peak Detection algorithm on the raw acceleration signal to detect steps ^34^. This algorithm demonstrated high accuracy (Mean 89%, SD 12%) during treadmill and outdoor walking ^34^. We calculated the step counts within each 60-second window and classified MPA, VPA and MVPA based on previously published step cadence thresholds (10–14). MPA was defined as a cadence ≥100 and <130 steps/min, VPA was defined as ≥ 130 steps/min, MVPA was defined as ≥ 100 steps/min. We considered a cadence of ≥ 20 and < 100 steps/min as light-PA (LPA). We used 20 steps/min as the lower cut-point for LPA to avoid misclassifying shuffling or pottering steps that occur in the 60-second window.

#### 2.2.3 Two-level cadence method

Accurate step detection is challenging in free-living environments. A study showed that routinely above 20% mean average percentage error (MAPE) existed in step counting accuracy in wrist-worn accelerometer cohort studies ^35^., likely due to the random wrist and upper limb movement (e.g., eating, brushing teeth, typing keyboard) misclassified as “steps”. To improve step counting accuracy, a hybrid two-level cadence method was developed (2LC). Firstly, we identified stepping activities (e.g., walking, running) by applying the RF classifier used in the ML method. Secondly, we summed up the step counts in 10-second windows classified as walking within each non-overlapping one minute interval, and classified that minute as LPA, MPA and VPA based on the same cadence thresholds applied in 1LC ^10^ . Then we added up the minutes corresponding to each intensity level, respectively, and calculated the mean daily duration across intensities by averaging over valid monitoring days.

### 2.3 Outcome ascertainment

Participants were followed up until 30 November 2022. The deaths records were obtained through linkage with the National Health Service (NHS) Digital of England and Wales or the NHS Central Register and National Records of Scotland. Based on the ICD-10 (International Classification of Diseases, 10^th^ revision) from both primary and contributory death cause, we defined CVD mortality as death from diseases of the circulatory system (ICD-10 codes: I0, I11, I13, I20–I51, I60–I69), excluding hypertension and diseases of arteries and lymph.

### 2.4 Covariates

We adjusted for covariates that are typical in studies examining intensity-specific PA and CVD risk based on previous peer-reviewed literature ^36^. We included age, sex, ethnicity, accelerometer wear duration, LPA minutes, MPA minutes, VPA minutes, smoking status, alcohol consumption, sleep duration, diet, screen time, education level, parental history of CVD and cancer, and medication use of cholesterol, blood pressure, or diabetes.

### 2.5 Statistical analysis

To minimize the influence of the sparse data, we excluded the MVPA, MPA, and VPA duration derived from the cadence-based and ML method above the 95th percentile ^4,6,7^. We examined the time-to-event dose-response association (hazard ratios [HRs]) of duration across intensities classified from all three methods with ACM and CVD mortality using the Cox proportional hazards regression with the 5th percentile of the duration across intensities as the reference and age as timescale. We demonstrated the dose-response association using the restricted cubic spline with knots placed at the 6th, 34th, and 67th percentile to fit the right-skewed distribution ^6^. We assessed the proportional hazards assumptions using Schoenfeld residuals and observed no violations. We presented overlapping plots to visually show and compare the dose-response associations of daily duration at each PA intensity with ACM and CVD mortality across the cadence-based and ML methods. To compare across methods, we assessed the minimum dose ^4,6,7^, 25th and 75th percentile dose (indicated by the ED50, ED25, ED75 in figures and tables, which estimate the daily duration associated with 50%, 25% and 75% of the optimal risk reduction, respectively), and the optimal dose (the nadir of the dose-response curve, representing the optimal risk reduction) and median daily duration and their corresponding HRs (95% CI). We used bootstrapping with replacement (1000 iterations) to calculate the 95% confidence interval for above metrics. We also assessed the dose-response association of the standardized MVPA/MPA/VPA duration as computed by each method (defined as absolute duration divided by the corresponding standard deviation [SD]) with the two mortality outcomes.

To assess the robustness of our results, we performed the sensitivity analysis on the 90th percentile of dataset derived from each method to further minimise the influence of sparse minutes, and on the participants based on the 95^th^ percentile of the 1LC method.

We performed all analyses using *R* software (version 4.2.2) and fitted models using *rms* package (version 6.3.0).

## 3. Results

### 3.1. Description of the study sample

**Supplementary Table 1** shows the characteristics of the sample derived by the 1LC method in MVPA minutes, stratified by quartiles of daily MVPA minutes. The mean (SD) age was 61.8 (7.8), and 39,847 (58.1%) were women. Over a mean follow-up 8.0 (0.9) years, there were 2134 deaths, of which 575 were due to CVD related causes. **Supplementary Fig. 1** shows the complete sample deprivation process.

### 3.2. Associations of daily MVPA with all-cause and CVD mortality

**Fig. 1** shows that the L-shaped dose-response curve of MVPA duration with ACM derived from the ML and 1LC method were similar, which the association decreased in a near-linear fashion up to a similar nadir point (optimal duration: ML, 32.0 (30.3, 33.9), HR, 0.63 (0.53, 0.75); 1LC, 31.9 (30.0, 34.7), HR, 0.60 (0.53, 0.69)) and then increased in a near-linear fashion with 95% CI largely overlapped. The L-shaped association derived from the 2LC method exhibited a more pronounced ACM risk reduction than other methods till reaching its nadir point. The median, minimum, 25^th^, 75^th^ and the optimal doses in 2LC were substantially smaller than those in the 1LC and ML method, with slightly higher corresponding ACM HRs, e.g., the minimum dose **(Supplementary Table 2)**: ML vs 1LC vs 2LC, 14.9 (95% CI: 14.6, 15.2) vs 11.1 (10.8, 11.4) vs 2.8 (2.6, 2.8), HR: 0.82 (95% CI: 0.76, 0.87) vs 0.80 (0.76, 0.85) vs 0.83 (0.78, 0.88). **Fig. 2** shows that the association derived from the 2LC method exhibited a more pronounced CVD risk reduction than other methods until its nadir point. ML method provided the lowest CVD HR across all the metrics among the methods, e.g., the ED75 dose, ML vs 2LC, 20.0 (19.0, 23.5) vs 5.6 (4.1, 9.9); HR: 0.60 (0.50, 0.71) vs 0.69 (0.58, 0.83) **(Supplementary Table 3)**. However, there was less evidence (wider 95% CI) for CVD mortality than ACM due to fewer CVD mortality events.

**Fig. 1.**
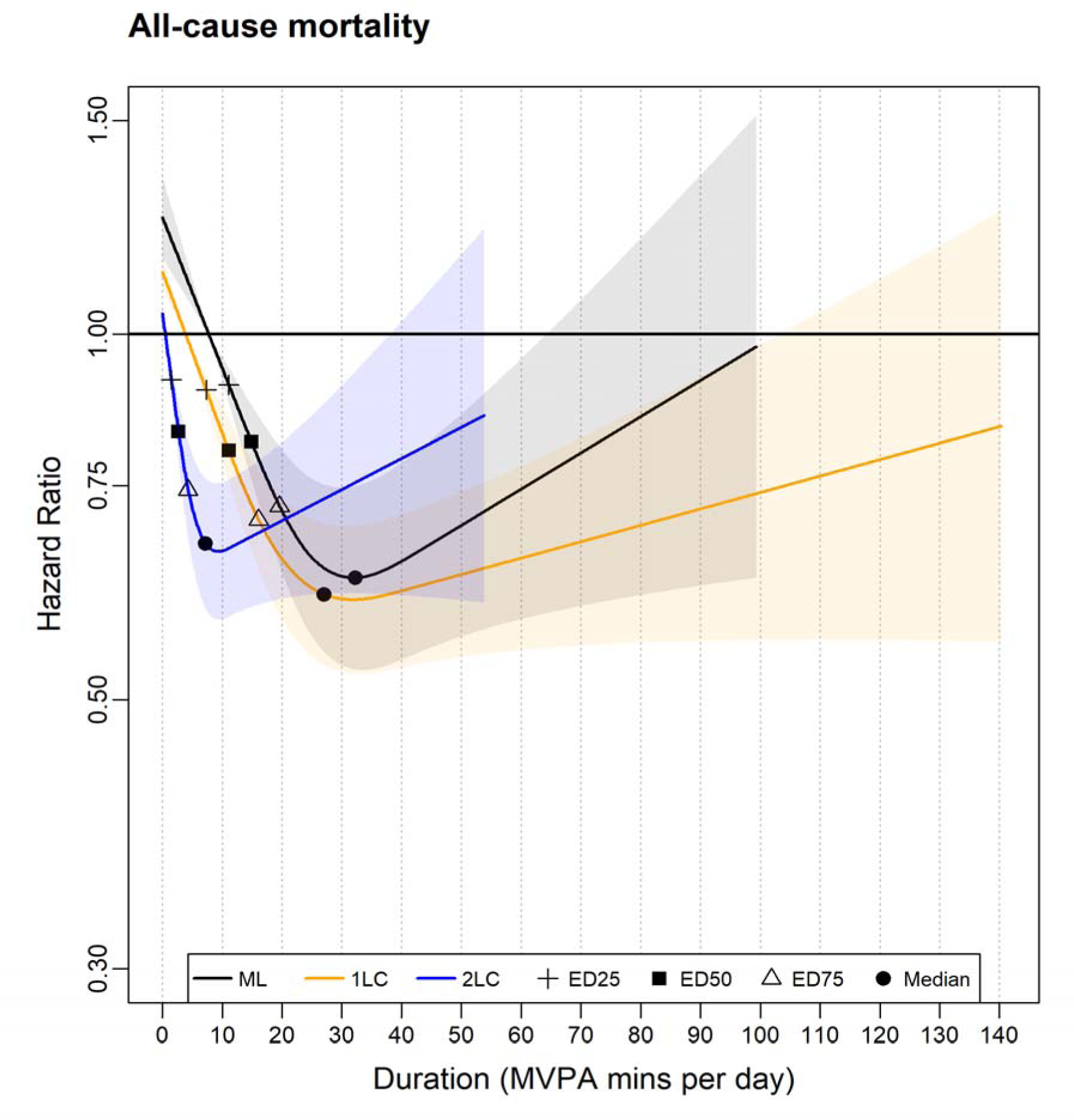
Association of the MVPA minutes derived from the cadence-based methods (1LC and 2LC) and machine learning (ML) method with all-cause mortality. 1LC indicates the one-level cadence method. 2LC indicates the two-level cadence method. Cross, diamond, and triangle represent the daily MVPA minutes that associated with 25% (ED25), 50% (ED50), and 75% (ED75) of the optimal risk reduction, respectively, and circle represents the median daily MVPA minutes. Please see supplementary table 2 for the list of metrics (duration and corresponding HR). The association was adjusted for age, sex, wear time, light physical activity duration, smoking status, alcohol consumption, sleep duration, diet, screen-time, education, self-reported parental history of CVD and cancer, and self-reported medication use (cholesterol, blood pressure, and diabetes). The range was capped at the 95th percentile to minimize the influence of sparse data. All methods used their corresponding 5th percentile as the reference level to calculate each HR.

**Fig 2.**
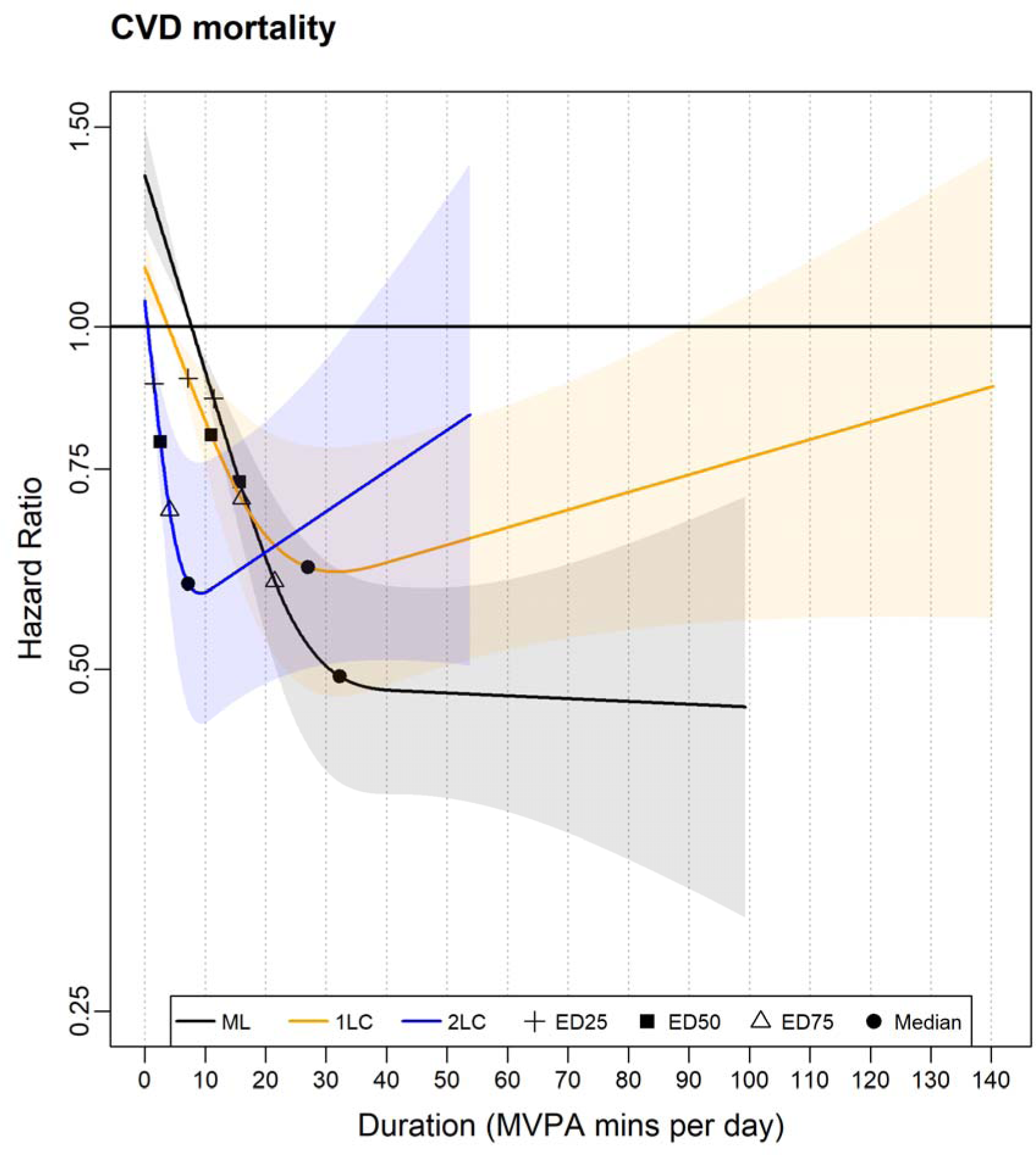
Association of the MVPA minutes derived from the cadence-based methods (1LC and 2LC) and machine learning (ML) method with CVD mortality. 1LC indicates the one-level cadence method. 2LC indicates the two-level cadence method. Cross, diamond, and triangle represent the daily MVPA minutes that associated with 25% (ED25), 50% (ED50), and 75% (ED75) of the optimal risk reduction, respectively, and circle represents the median daily MVPA minutes. Please see supplementary table 2 for the list of metrics (duration and corresponding HR). The association was adjusted for age, sex, wear time, Light PA, smoking status, alcohol consumption, sleep duration, diet, screen-time, education, self-reported parental history of CVD and cancer, and self-reported medication use (cholesterol, blood pressure, and diabetes). The range was capped at the 95th percentile to minimize the influence of sparse data. All methods used their corresponding 5th percentile as the reference level to calculate each HR.

### 3.3. Associations of MPA duration with all-cause and CVD mortality

**Fig. 3** shows that the L-shaped dose-response association of MPA mins between the ML and 1LC method with ACM were similar with 95% CI largely overlapped, and both methods exhibited linear inverse association up to around the median duration (ML:32.2 [32.1, 32.3], HR, 0.60 [0.51, 0.70]; 1LC: 19.9 [19.9, 20.0], HR, 0.65 [0.58, 0.73]) (**Supplementary Table 2)** where the curve flattened. Meanwhile, the 2LC method exhibited steeper risk reduction in a near-linear fashion up to its median (5.3 [5.3, 5.4], HR, 0.68 [0.60, 0.77]) and continued to show more risk reduction than other methods until 20 MPA minutes. The ML method provided substantially larger minimum, 25^th^, and 75^th^ percentile durations than the 1LC and 2LC methods, with more ACM risk reduction, e.g., the minimum dose (**Supplementary Table 2)**: ML vs 1LC vs 2LC, 14.9 (95% CI: 14.6, 15.2) vs 8.6 (8.3, 9.3) vs 2.3 (2.1, 2.8). HR: 0.77 (0.70, 0.83) vs 0.81 (0.76, 0.86) vs 0.82 (0.77, 0.88). **Fig. 4** shows that L-shaped beneficial association with CVD mortality across methods, respectively, was similar to that in the MPA-ACM association in terms of the dose-response curve and other metrics **(Supplementary Table 3),** except for the upward pattern of the 2LC method after the median MPA mins (roughly 6 mins).

**Fig. 3.**
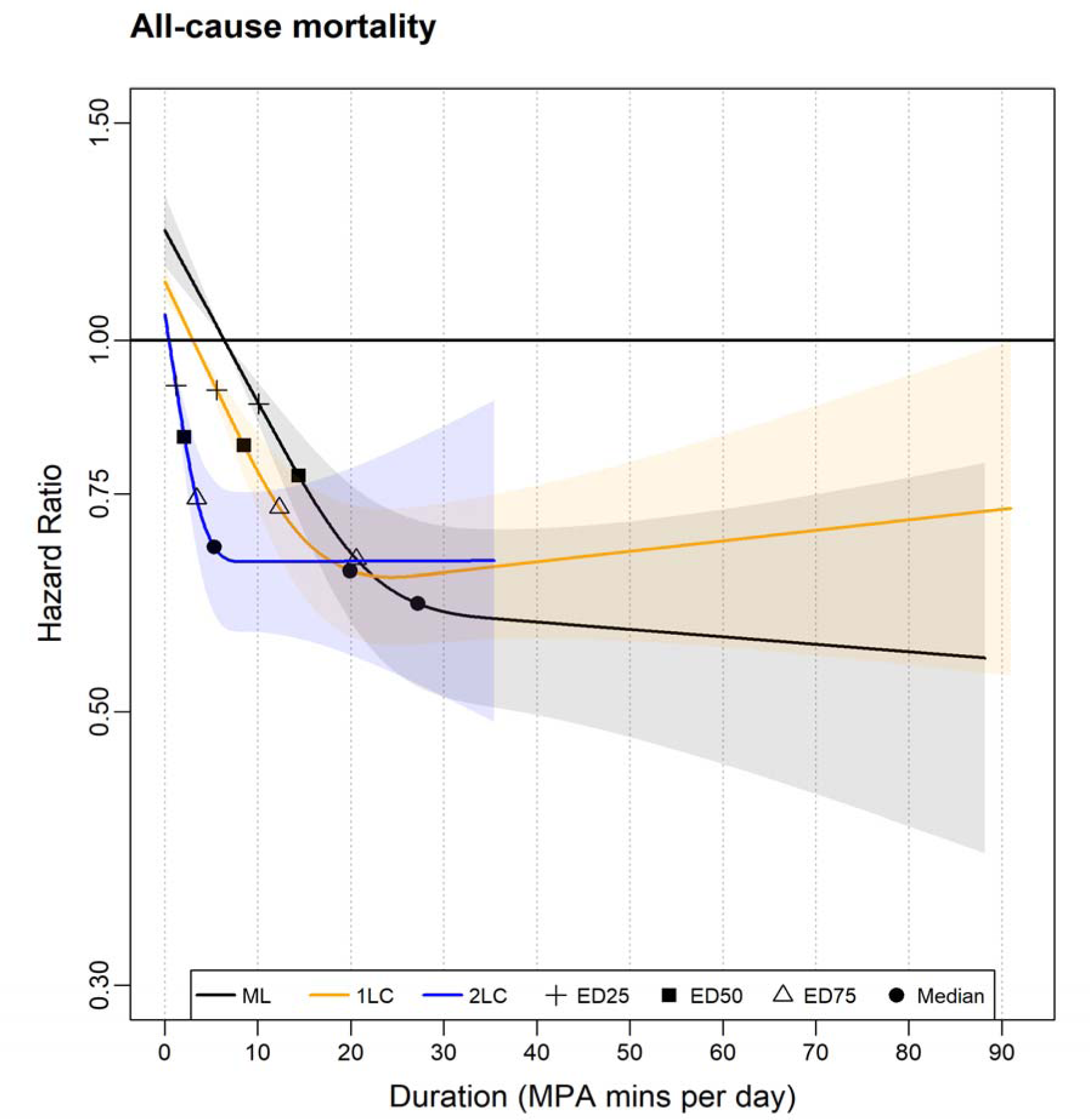
Association of the MPA minutes derived from the cadence-based methods (1LC and 2LC) and machine learning (ML) method with all-cause mortality. 1LC indicates the one-level cadence method. 2LC indicates the two-level cadence method. Cross, diamond, and triangle represent the daily MVPA minutes that associated with 25% (ED25), 50% (ED50), and 75% (ED75) of the optimal risk reduction, respectively, and circle represents the median daily MVPA minutes. Please see supplementary table 2 for the list of metrics (duration and corresponding HR). The association was adjusted for age, sex, wear time, light PA, VPA, smoking status, alcohol consumption, sleep duration, diet, screen-time, education, self-reported parental history of CVD and cancer, and self-reported medication use (cholesterol, blood pressure, and diabetes). The range was capped at the 95^th^ percentile to minimize the influence of sparse data. All methods used their corresponding 5th percentile as the reference level to calculate each HR.

**Fig 4.**
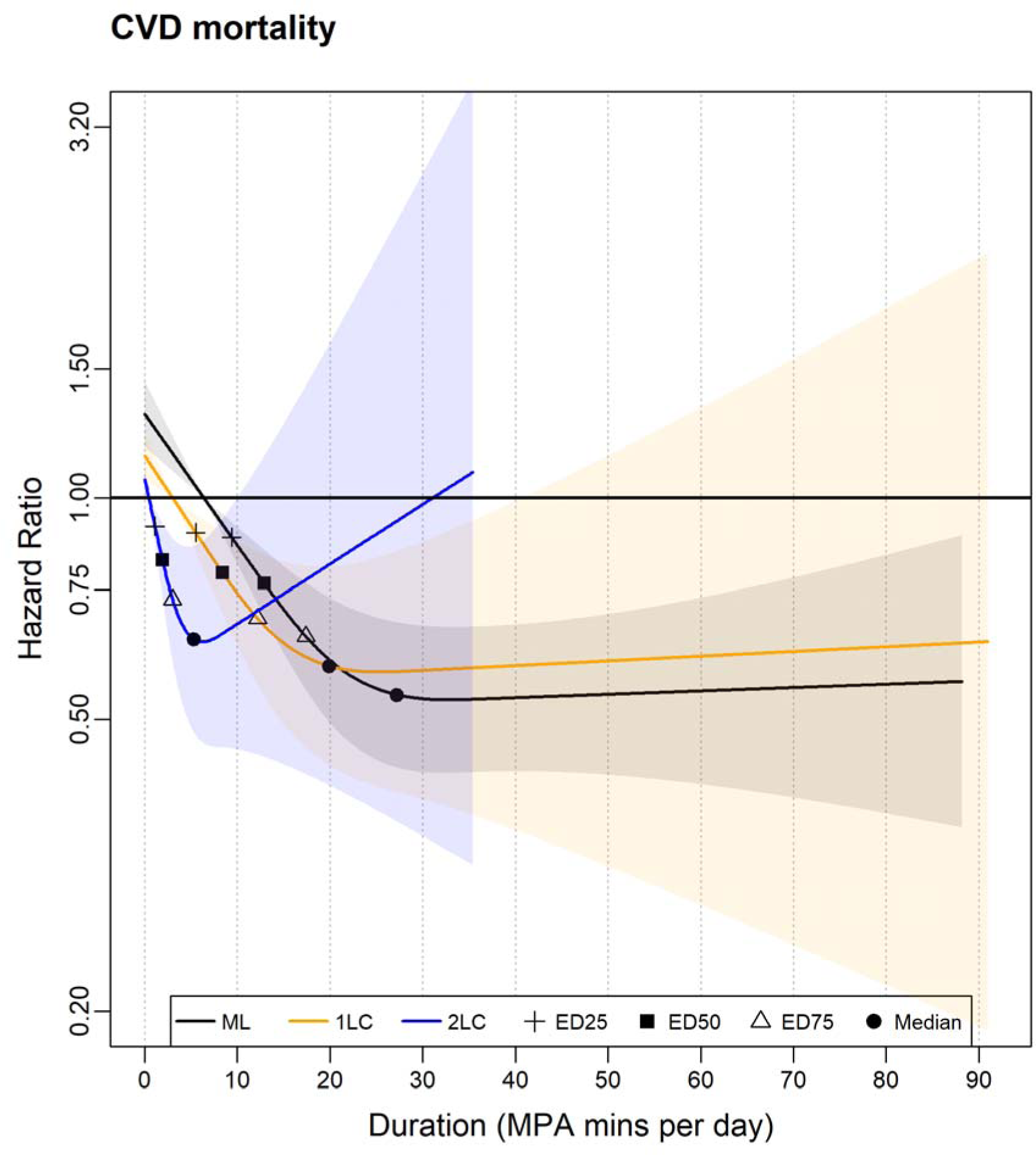
Association of the MPA minutes derived from the cadence-based methods (1LC and 2LC)* and machine learning (ML) method with CVD mortality. 1LC indicates the one-level cadence method. 2LC indicates the two-level cadence method. Cross, diamond, and triangle represent the daily MVPA minutes that associated with 25% (ED25), 50% (ED25), and 75% (ED25) of the optimal risk reduction, respectively, and circle represents the median daily MVPA minutes. Please see supplementary table 2 for the list of metrics (duration and corresponding HR). The association was adjusted for age, sex, wear time, light PA, VPA, smoking status, alcohol consumption, sleep duration, diet, screen-time, education, self-reported parental history of CVD and cancer, and self-reported medication use (cholesterol, blood pressure, and diabetes). The range was capped at the 95th percentile to minimize the influence of sparse data. All methods used their corresponding 5th percentile as the reference level to calculate each HR.

### 3.4. Associations of VPA duration with all-cause and CVD mortality

**Fig. 5** shows that the 1LC method provided the least pronounced dose-response association of VPA mins with ACM, and the ML method provided the most pronounced one, e.g., the median **(Supplementary Table 2)**: ML, 3.2 (95% CI: 3.2, 3.2), HR, 0.53 (95% CI: 0.44, 0.64); 1LC, 6.9 (6.9, 7.0), 0.68 (0.60, 0.77); 2LC, 2.0 (2.0, 2.0), 0.69 (0.61, 0.79). The dose-response association of the 2LC method was almost identical to that of the ML method up to roughly 1.5 mins, with both methods showing steep risk reduction. The ML method further exhibited steep ACM risk reduction up to around its median point and demonstrated substantially lower ACM risk reduction than that of the 2LC method. **Fig. 6** shows that the dose-response association of VPA minutes derived by the 2LC method with CVD mortality exhibited more pronounced risk reduction compared to the ML method up to around 10 minutes where the curve flattened. The 2LC method provided smaller dose but higher corresponding CVD HRs for most point estimates, e.g, the minimum point **(Supplementary Table 3):** ML: 9.0 (7.4, 9.3), HR, 0.70 (0.60, 0.81); 2LC, 0.6 (0.4, 0.9), HR, 0.75 (0.67, 0.85). However, there was less evidence (wider 95% CI) for CVD mortality than ACM due to fewer CVD mortality events.

**Fig. 5.**
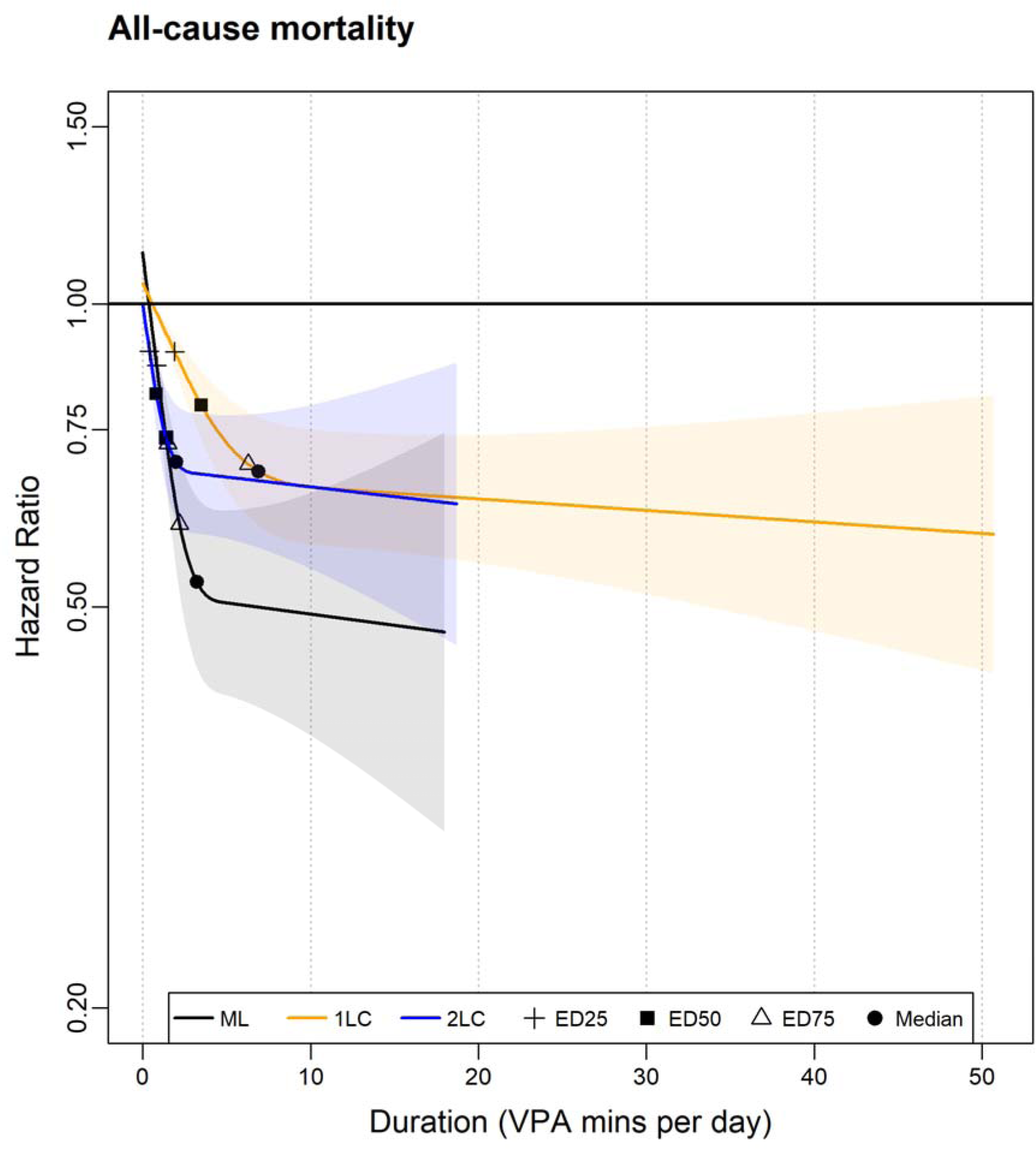
Association of the VPA minutes derived from the cadence-based methods (1LC and 2LC) and machine learning (ML) method with all-cause mortality. 1LC indicates the one-level cadence method. 2LC indicates the two-level cadence method. Cross, diamond, and triangle represent the daily MVPA minutes that associated with 25%(ED25), 50% (ED50), and 75% (ED75) of the optimal risk reduction, respectively, and circle represents the median daily MVPA minutes. Please see supplementary table 2 for the list of metrics (duration and corresponding HR). Adjusted for age, sex, wear time, light PA, MPA, smoking status, alcohol consumption, sleep duration, diet, screen-time, education, self-reported parental history of CVD and cancer, and self-reported medication use (cholesterol, blood pressure, and diabetes). The range was capped at the 95th percentile to minimize the influence of sparse data. All methods used their corresponding 5th percentile as the reference level to calculate each HR.

**Fig 6.**
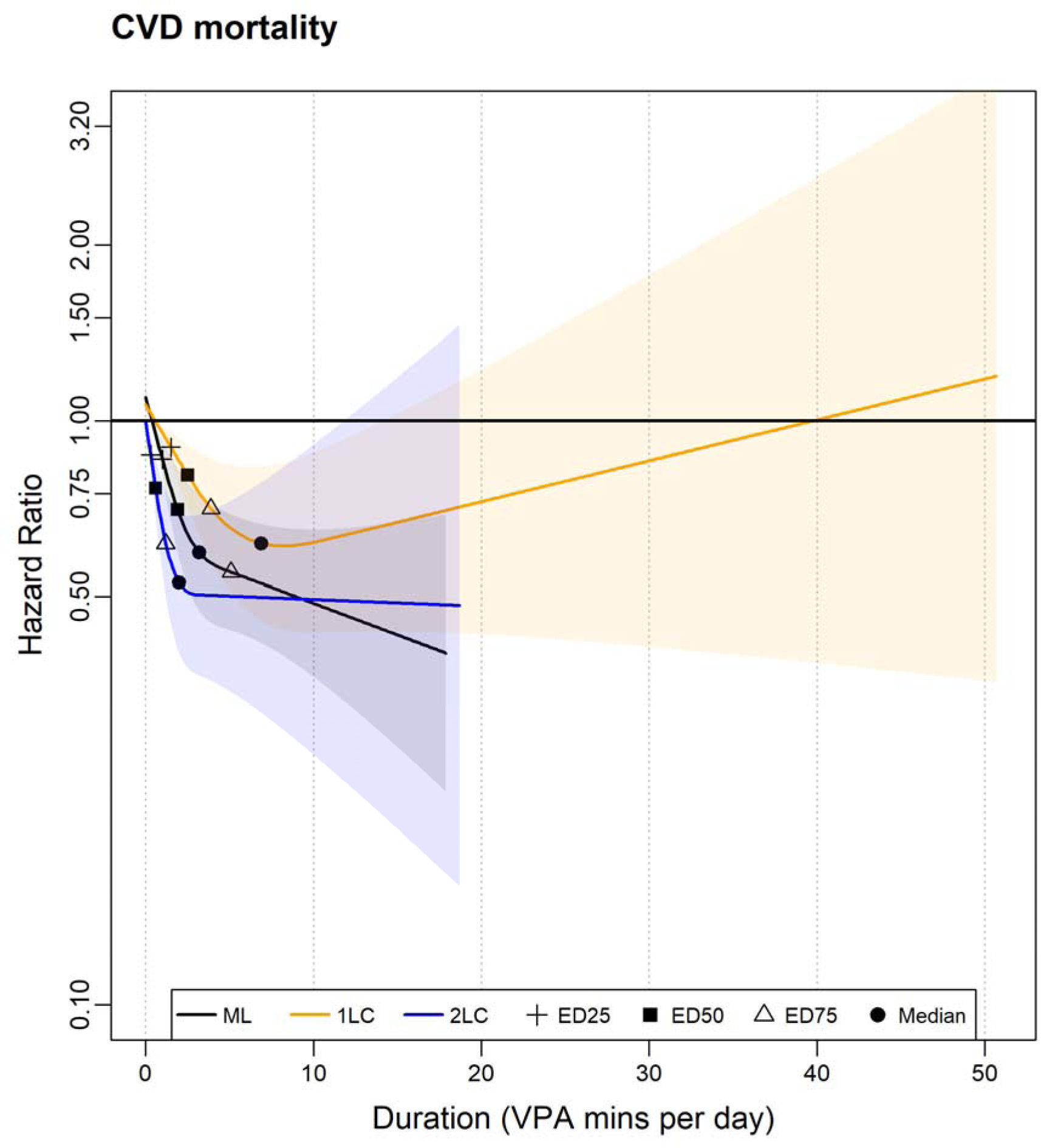
Association of the VPA minutes derived from the cadence-based methods (1LC and 2LC)* and machine learning (ML) method with CVD mortality. 1LC indicates the one-level cadence method. 2LC indicates the two-level cadence method. Cross, diamond, and triangle represent the daily MVPA minutes that associated with 25% (ED25), 50% (ED50), and 75% (ED75) of the optimal risk reduction, respectively, and circle represents the median daily MVPA minutes. Please see supplementary table 2 for the list of metrics (duration and corresponding HR). The association was adjusted for age, sex, wear time, light PA, MPA, smoking status, alcohol consumption, sleep duration, diet, screen-time, education, self-reported parental history of CVD and cancer, and self-reported medication use (cholesterol, blood pressure, and diabetes). The range was capped at the 95th percentile to minimize the influence of sparse data. All methods used their corresponding 5th percentile as the reference level to calculate each HR.

### 3.5. Associations of the standardised duration with all-cause and CVD mortality

**Supplementary Fig 2-4** show that the dose-response associations of standardized duration with ACM across intensities, respectively, was similar between the 1LC and 2LC method, with 95% CI largely overlapped. The ML method provided the most pronounced association of standardized duration with ACM across intensities, especially for the standardized VPA minutes-ACM association (**Supplementary Fig 4),** e.g., for the median of the standardized VPA minutes/day and the corresponding HR (ML: -0.31, HR, 0.53 (0.44, 0.64); 1LC: -0.35, HR, 0.68 (0.60, 0.77); 2LC: -0.39, HR, 0.71, (0.63, 0.81)) (**Supplementary Table 4)**. **Supplementary Table 4** shows that almost all the metrics related to MPA and VPA between the 2LC and the ML method were different. **Supplementary Fig 5 and 7** show that the 95% CI of the L-shaped dose-response association of the standardized MVPA and VPA duration with CVD mortality were largely overlapped. **Supplementary Fig 6** shows that the ML method provided the most pronounced association of standardized MPA duration with CVD mortality compared to other methods, e.g., for the median of the standardized MPA minutes/day and the corresponding HR (ML: -0.20, HR, 0.54 (0.43,0.68); 1LC: -0.27, HR, 0.59 (0.43,0.81); 2LC: -0.35, HR, 0.64 (0.48, 0.86)) (**Supplementary Table 5)**. We observed less evidence (wider 95% CI) for association of standardized duration with CVD mortality due to fewer CVD mortality events than ACM.

## 4. Discussion

To our knowledge, this is the first study to compare the dose-response associations of different PA intensity classification methods (cadence-based methods and ML method) with standard health outcomes used routinely in observational studies. The ML and 1LC methods provided similar dose-response associations. The 2LC method exhibited the most pronounced rate of decrease in the association with mortality across intensities at the lower end of the duration. The ML elicited more risk reduction in mortality than cadence-based methods, particularly for the VPA-ACM association. The ML method provided the most pronounced dose-response associations of the standardized duration with mortality outcomes across different intensity levels. Future research could focus on deriving the cadence-thresholds for short windows, e.g., 10-second window for each PA intensity level, to inform the step-based public health guideline and interventions, etc.

A potential explanation for the variation in dose-response associations is the different window length applied in the 2LC and ML method. Our ML method classified PA intensity using 10-second window, enabling the detection of finer pattern of MVPA derived by very brief bouts, thus leading to different dose-response association relative to the 2LC method which applied 60-second window. The shorter window is more likely to detect MVPA burst ^37^. Prior studies applying various window lengths (15 seconds, 30 seconds, 60 seconds) demonstrated that shorter windows detect significantly more MVPA minutes ^27,38^. Applying a longer epoch such as 60-seconds window might result in a smoothing effect ^39^, e.g., a 30-second jogging to catch bus (MVPA burst) might be degraded to lower intensity using 2LC method. A recently published large cohort study has indicated that the MVPA accumulated through bouts that less than 1 min (with VPA being at least 15% proportion of the MVPA time) would elicit beneficial change on ACM risk ^5^. Such smoothing effect cause higher intensity PA to decrease to a lower level, potentially misleading researchers to unreliable intensity time – morality association and therefore overlooked the health effect of the valuable granularity of intensity time.

Another potential explanation is that the 2LC method, although be able to detect stepping activities and thus counting steps accurately, might overlook the variation in step intensity (e.g., steps involving upper-body movements). The ML method, in conjunction with the wrist accelerometer, could detect steps associated with upper-body movements ^40^ (e.g., ambulatory gardening), thus capturing wider range of PAs and providing more representative duration of PA intensities than 2LC method. Prior studies have indicated that PAs involving upper-body movements such as occupational and household activities account for a large proportion of MVPA time (e.g., the household PAs was the most prevalent domain for those who reported under weekly 150 MVPA mins) ^41^.

The association of the VPA duration derived by the ML method with ACM was similar to that of 2LC method at the lower end of distribution (up to 2 mins), suggesting that these VPA duration might be the result of incidental brisk walking or running (e.g., quickly moving for 10 seconds). Prior study has shown that a large portion (92.3%) of VILPA bouts in non-exercisers were less than a minute ^4^. The 2LC using 60-seconds window may overlook many such bursts and thus unlikely to derive the dose-response association close to ML method in free-living environment. The ML-derived VPA dose-response association appeared the steepest HR reduction for ACM with the lowest HR associated in all point estimates, suggesting that the VPA duration including granularity duration might demonstrate more beneficial change on ACM than that of the VPA duration calculated based on step counts per min. This enhanced the value of the short VPA burst for achieving beneficial health change.

After standardization of the MVPA/MPA/VPA minutes, the dose-response association with mortality between the 2LC and the ML method showed variation in terms of the curve and most point estimates. This was especially evident in the association of standardized VPA duration with the ACM. This finding implied that the propensity of the estimation methods not only provide different estimates of the duration, but also the additional duration of the MVPA/MPA/VPA bursts detected by the ML method elicited higher degree of mortality risk reduction than other methods. This enhanced the role of the short bout higher intensity PA in potentially beneficial changes on health.

Wrist, as the most active part of body, produces many extraneous wrist movements (e.g., eating) which might be recorded as ambulatory by wrist-worn accelerometers (“error steps”) ^42^. Although the 1LC method was based on the same cadence-thresholds as 2LC, it counted steps without identifying the stepping activities first, which leads to substantial higher “error steps” counts than the 2LC method. Therefore, the 1LC method inflated MVPA/MPA durations and thus derived similar association to the ML method. A study comparing the step detection accuracy of 13 wearables found that wrist-worn accelerometers detected roughly 450 more steps per day than waist-worn in free-living environment ^43^. The inflated MVPA/MPA duration might lead to associations with mortality close to that of ML method. Nonetheless, the inherent randomness in wrist movements might introduce variations in error steps count, influenced by factors such as the variation of wrist-movements, sample characteristics or step-detection methods making 1LC-derived dose-response association with mortality unstable.

To our knowledge, this is the first study comparing the PA intensity classification methods (cadence-based and ML method) in terms of the association with standard health outcomes used routinely in observational studies. Our study had large sample with long follow-up years. The ML method could detect the micro-pattern of PA and upper-body movements in free-living environment. The 2LC method used advanced step detection algorithm and provide better accuracy. We have conducted multiple sensitivity analyses to show robustness of our findings. Our study had several limitations. Our study categorized confounders (e.g., sleep duration, LPA) to avoid statistical assumption violation. The ML method is not the ground-truth PA intensity time estimation method, although showed high accuracy for MPA (precision: 92.7) and VPA (94.6) comparing to the criterion data^7^. The measurement error in the ML method for PA intensity time might affect the comparison of the intensity time - mortality association among methods. The UK biobank has a low response-rate (5.5%) and thus not representative of the population ^44^, although a study has demonstrated that poor representativeness in UK biobank didn’t materially affect the PA-mortalities associations ^45^.

## 5. Perspective

The 1LC method produced similar dose-response MVPA/MPA-mortality associations to the ML method. The 2LC method exhibited the most pronounced associations with mortality across intensities than the ML method at lower end of the duration. The ML method elicited more risk reduction in mortality than cadence-based methods, particularly for the VPA-ACM association. After standardization of the duration, the ML method generally provided most pronounced associations across intensities. These suggested that the MPA/VPA bursts detected by the shorter detection window (10-second) in the ML method can capture the associations between activities and health outcomes with greater granularity. The method that based on the threshold of walking steps in a minute long window might underestimate both the duration and potential health benefits of the MPA and VPA bouts under the free-living conditions. Future studies could focus on the cadence-based thresholds for windows less than a minute for detecting the MPA and VPA bursts.

## Acknowledgements

This research has been conducted using the UK Biobank Resource under Application Number 25813. The authors would like to thank all the participants and professionals contributing to the UK Biobank.

## Authors’ contributions

E.S., M.N.A, L.W., R.K.B, and S.G.T contributed to the study conception and design, and interpretation of data. Data preparation was performed by M.N.A. The draft of the manuscript and data analysis were performed by L.W. All authors revised it critically and gave final approval and agreed to be accountable for all aspects of the work, ensuring integrity and accuracy.

## Informed Consent

Informed consent was obtained from all individual participants included in the study.

## Data Availability Statement

This research has been conducted using the UK Biobank Resource under Application Number 25813. Bona fide researchers can register and apply to use the UK Biobank dataset at http://ukbiobank.ac.uk/register-apply/.

---

**Supplemental fig 1.**
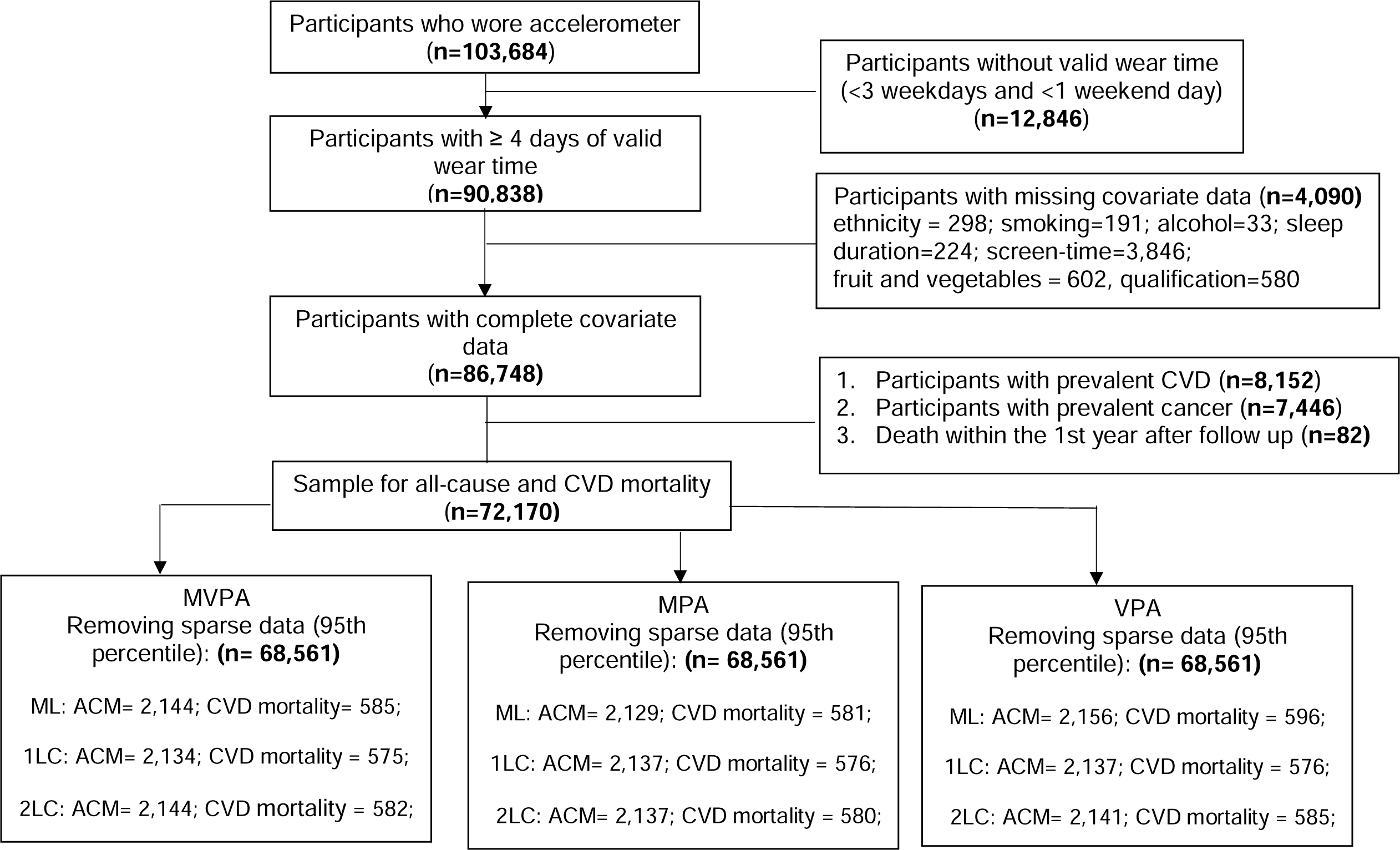
Flow diagram of participants.

**Supplementary fig 2.**
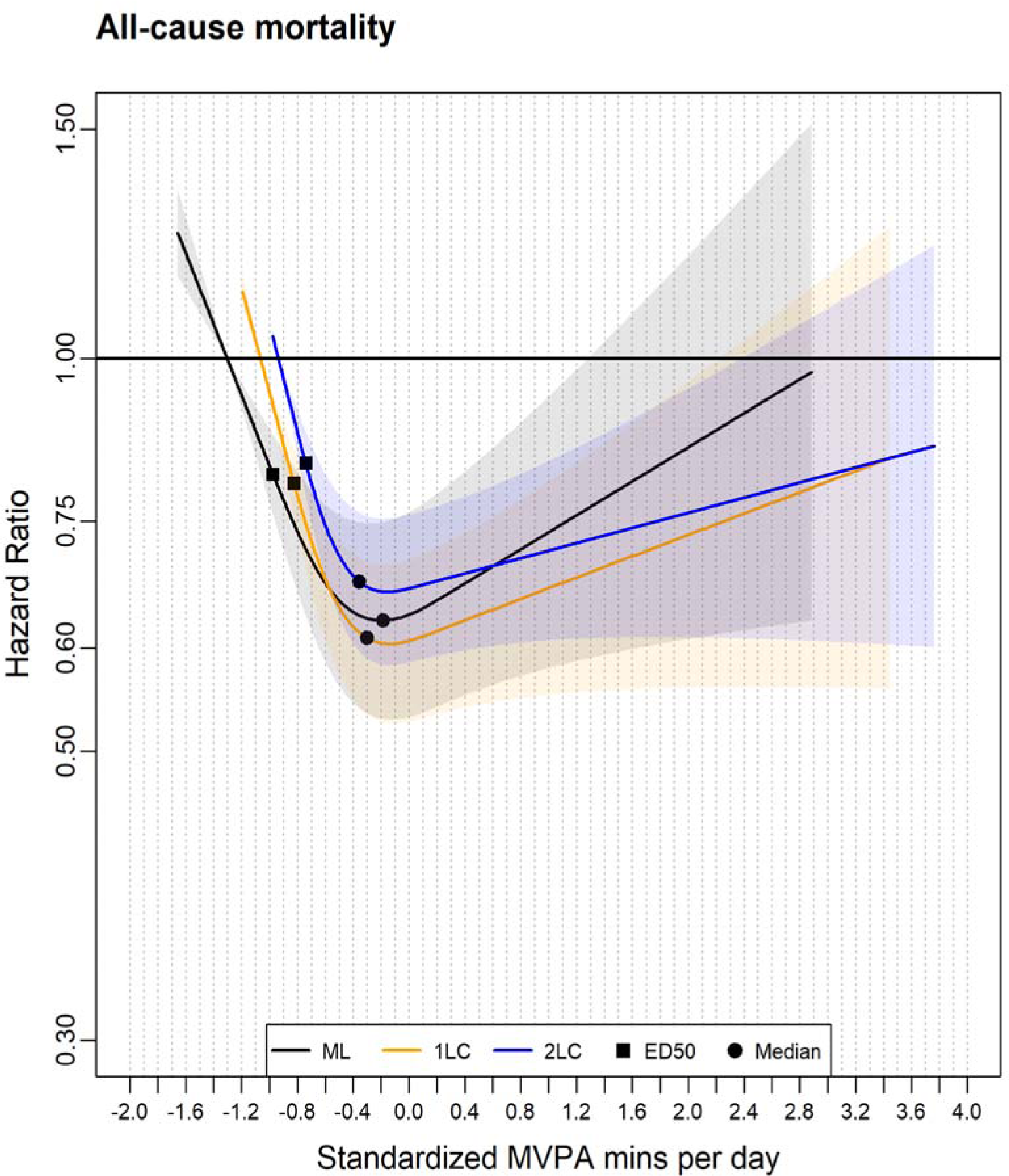
Association of the standardized MVPA minutes derived from the cadence-based methods (1LC and 2LC) and machine learning (ML) method with all-cause mortality. 1LC indicates the one-level cadence method. 2LC indicates the two-level cadence method. Diamond represents the minimum dose (ED50), which estimates the daily standardised MVPA minutes associated with 50% of optimal risk reduction across methods. Circle represents the standardised daily median MVPA minutes across methods. Please see supplementary table 3 for the list of metrics (duration and corresponding HR). The association was adjusted for age, sex, wear time, LPA, smoking status, alcohol consumption, sleep duration, diet, screen-time, education, self-reported parental history of CVD and cancer, and self-reported medication use (cholesterol, blood pressure, and diabetes). The range was capped at the 95th percentile to minimize the influence of sparse data. All methods used their corresponding 5th percentile as the reference level to calculate each HR.

**Supplementary fig 3.**
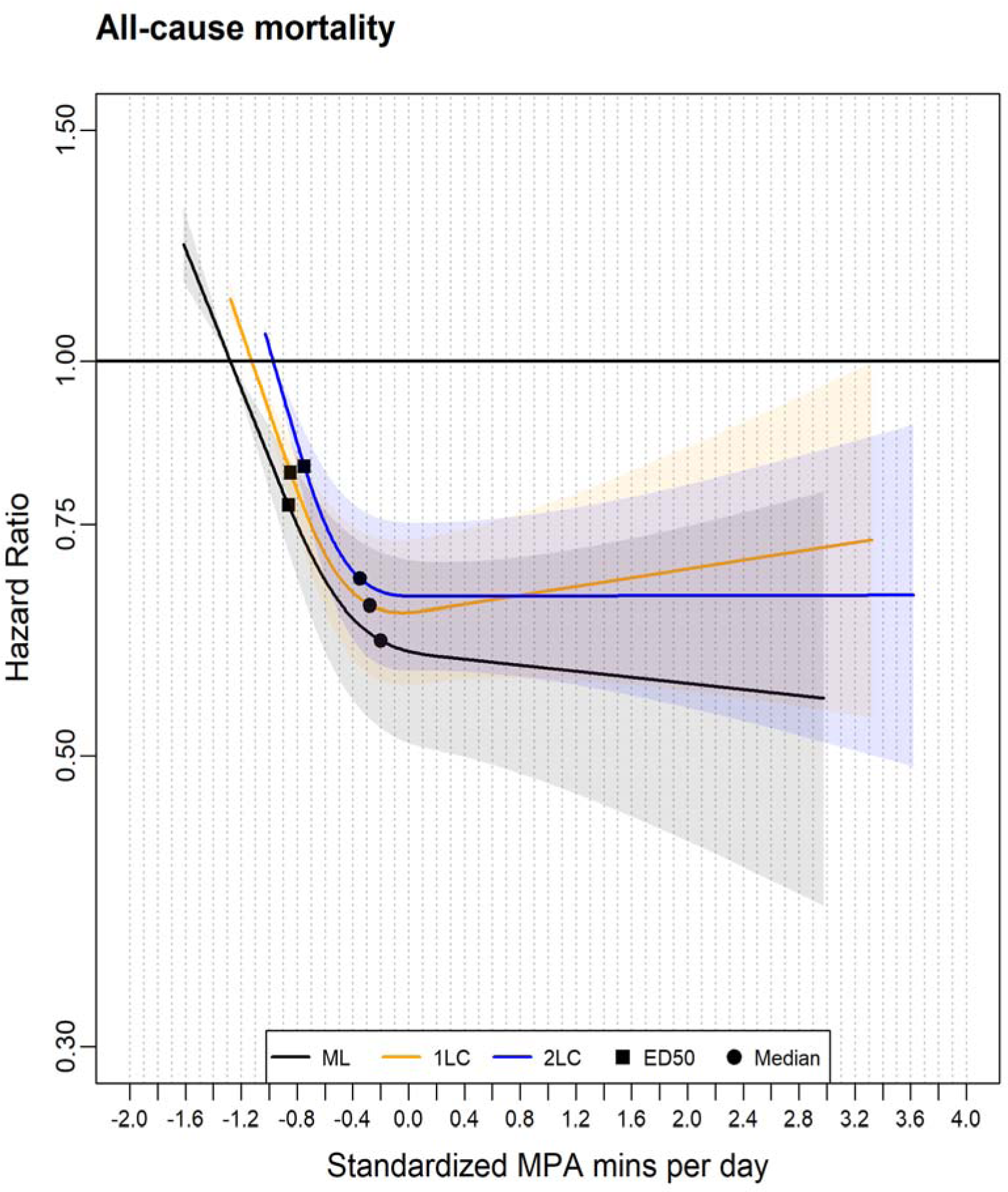
Association of the standardized MPA minutes derived from the cadence-based methods (1LC and 2LC)* and machine learning (ML) method with All-cause mortality. 1LC indicates the one-level cadence method. 2LC indicates the two-level cadence method. Diamond represents the minimum dose (ED50), which estimates the daily standardised MPA minutes associated with 50% of optimal risk reduction across methods. Circle represents the standardised median MPA minutes across methods. Please see supplementary table 3 for the list of metrics (duration and corresponding HR). The association was adjusted for age, sex, wear time, LPA, VPA, smoking status, alcohol consumption, sleep duration, diet, screen-time, education, self-reported parental history of CVD and cancer, and self-reported medication use (cholesterol, blood pressure, and diabetes). The range was capped at the 95^th^ percentile to minimize the influence of sparse data. All methods used their corresponding 5th percentile as the reference level to calculate each HR.

**Supplementary fig 4.**
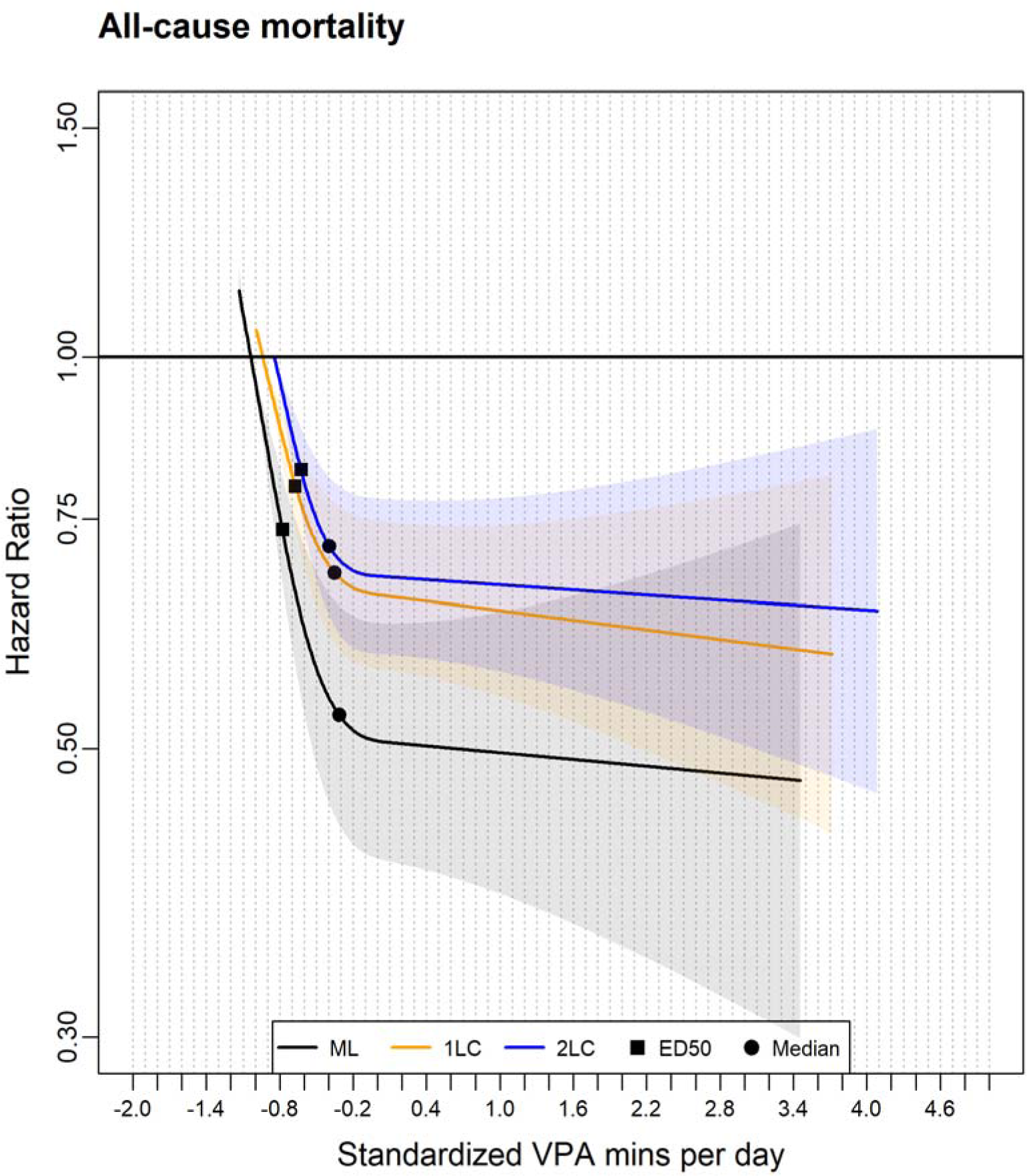
Association of the standardized VPA minutes derived from the cadence-based methods (1LC and 2LC) and machine learning (ML) method with all-cause mortality. 1LC indicates the one-level cadence method. 2LC indicates the two-level cadence method. Diamond represents the minimum dose (ED50), which estimates the daily standardised VPA minutes associated with 50% of optimal risk reduction across methods. Circle represents the standardised median VPA minutes across methods. Please see supplementary table 3 for the list of metrics (duration and corresponding HR). The association was adjusted for age, sex, wear time, LPA, MPA, smoking status, alcohol consumption, sleep duration, diet, screen-time, education, self-reported parental history of CVD and cancer, and self-reported medication use (cholesterol, blood pressure, and diabetes). The range was capped at the 95th percentile to minimize the influence of sparse data. All methods used their corresponding 5th percentile as the reference level to calculate each HR.

**Supplementary fig 5.**
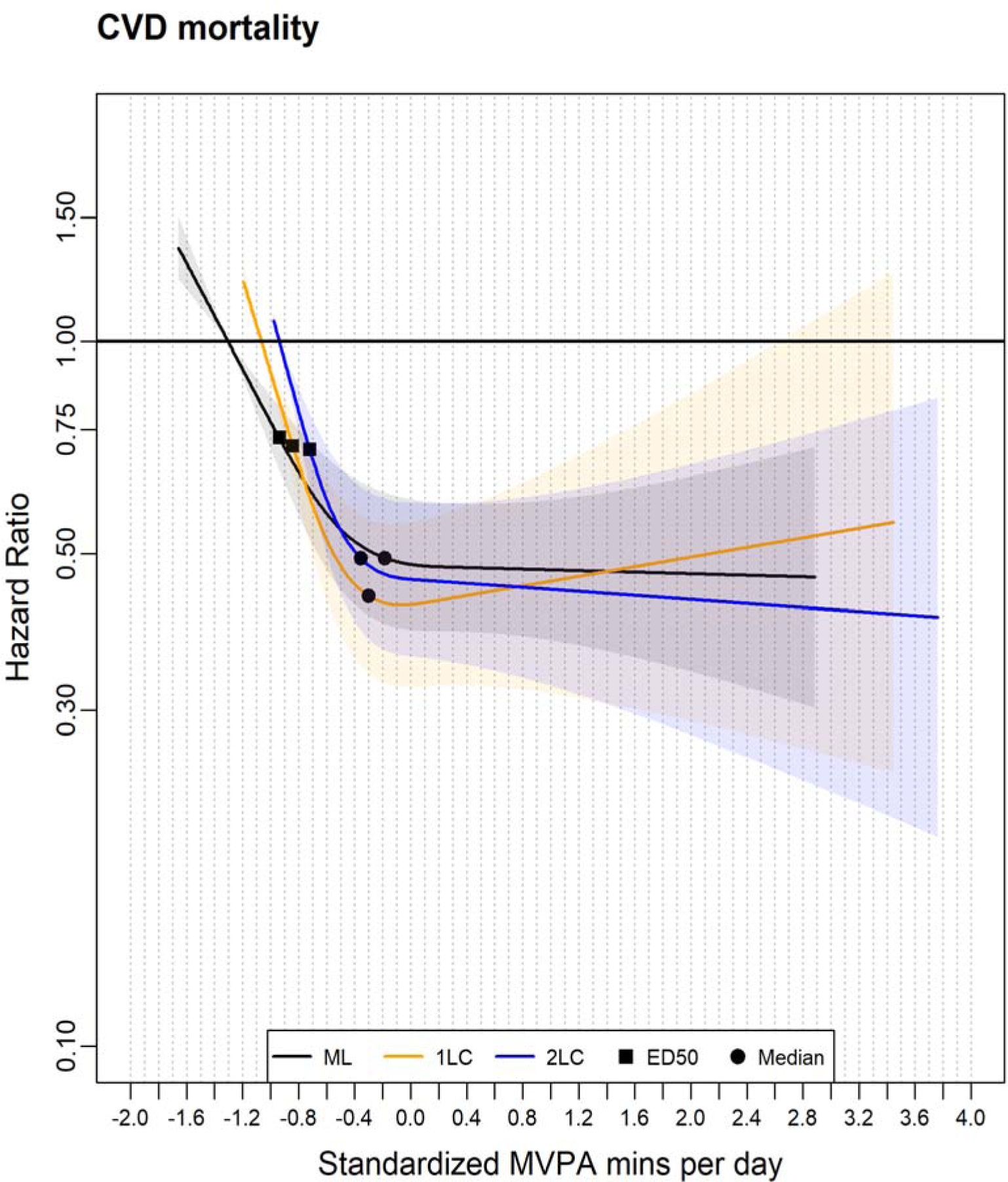
Association of the standardized MVPA minutes derived from the cadence-based methods (1LC and 2LC) and machine learning (ML) method with CVD mortality. 1LC indicates the one-level cadence method. 2LC indicates the two-level cadence method. Diamond represents the minimum dose (ED50), which estimates the daily standardised MVPA minutes associated with 50% of optimal risk reduction across methods. Circle represents the standardised median MVPA minutes across methods. Please see supplementary table 4 for the list of metrics (duration and corresponding HR). The association was adjusted for age, sex, wear time, LPA, smoking status, alcohol consumption, sleep duration, diet, screen-time, education, self-reported parental history of CVD and cancer, and self-reported medication use (cholesterol, blood pressure, and diabetes). The range was capped at the 95th percentile to minimize the influence of sparse data. All methods used their corresponding 5th percentile as the reference level to calculate each HR.

**Supplementary fig 6.**
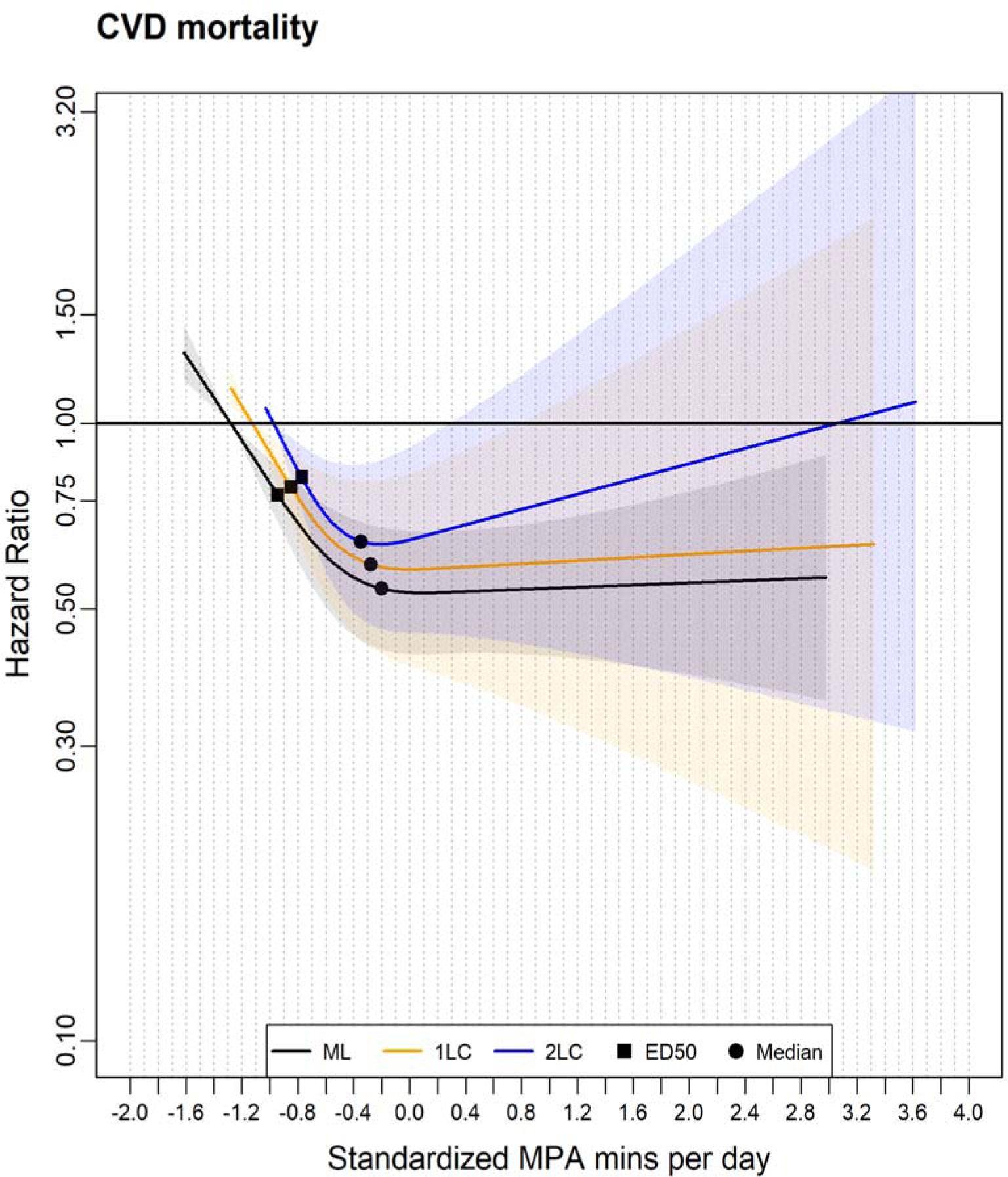
Association of the standardized MPA minutes derived from the cadence-based methods (1LC and 2LC) and machine learning (ML) method with CVD mortality. 1LC indicates the one-level cadence method. 2LC indicates the two-level cadence method. Diamond represents the minimum dose (ED50), which estimates the daily standardised MPA minutes associated with 50% of optimal risk reduction across methods. Circle represents the standardised median MPA minutes across methods. Please see supplementary table 4 for the list of metrics (duration and corresponding HR). The association was adjusted for age, sex, wear time, LPA, VPA, smoking status, alcohol consumption, sleep duration, diet, screen-time, education, self-reported parental history of CVD and cancer, and self-reported medication use (cholesterol, blood pressure, and diabetes). The range was capped at the 95th percentile to minimize the influence of sparse data. All methods used their corresponding 5th percentile as the reference level to calculate each HR.

**Supplementary fig 7.**
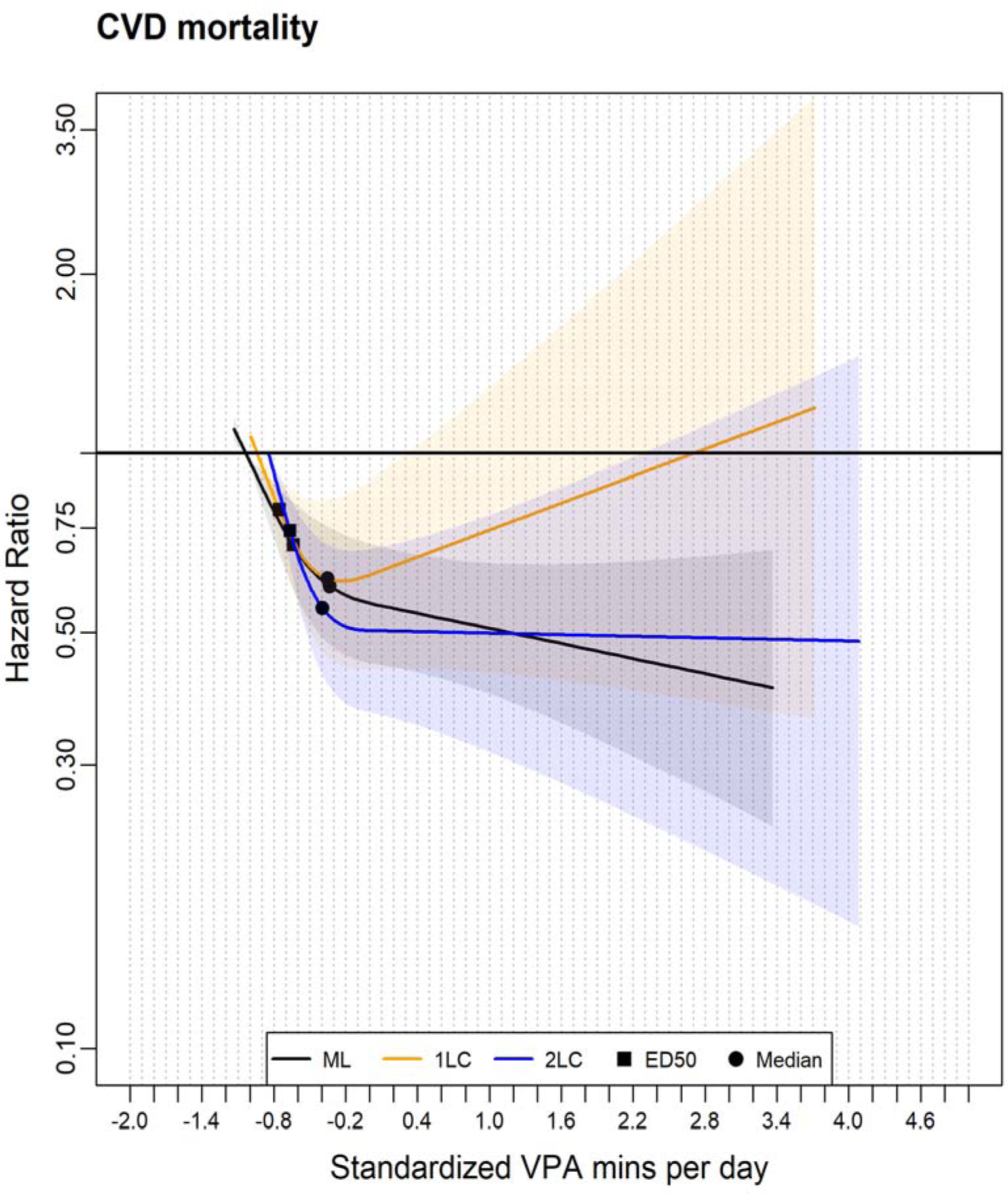
Association of the standardized VPA minutes derived from the cadence-based methods (1LC and 2LC) and machine learning (ML) method with CVD mortality. 1LC indicates the one-level cadence method. 2LC indicates the two-level cadence method. Diamond represents the minimum dose (ED50), which estimates the daily standardised VPA minutes associated with 50% of optimal risk reduction across methods. Circle represents the standardised median VPA minutes across methods. Please see supplementary table 4 for the list of metrics (duration and corresponding HR). The association was adjusted for age, sex, wear time, LPA, MPA, smoking status, alcohol consumption, sleep duration, diet, screen-time, education, self-reported parental history of CVD and cancer, and self-reported medication use (cholesterol, blood pressure, and diabetes). The range was capped at the 95th percentile to minimize the influence of sparse data. All methods used their corresponding 5th percentile as the reference level to calculate each HR.

**Supplementary Table 1.** Characteristics of Participants, by quartile a of one-level cadence method-derived MVPA minutes.

|  |  | **One-level cadence method derived daily MVPA**<br>**minutes** |  |  |  |
| --- | --- | --- | --- | --- | --- |
|  | **Total** | **Quartile 1** | **Quartile 2** | **Quartile 3** | **Quartile 4** |
| Total participants, n | 68561 | 17186 | 17097 | 17138 | 17140 |
| Male, n (%) | 28714<br>(41.9) | 6406 (37.3) | 7011 (41.0) | 7409 (43.2) | 7888 (46.0) |
| wear_days, mean (sd) | 6.8 (0.4) | 6.8 (0.4) | 6.8 (0.34) | 6.8 (0.4) | 6.8 (0.3) |
| follow-up year, mean (SD) | 8.0 (0.9) | 7.9 (1.0) | 8.0 (0.9) | 8.0 (0.8) | 8.0 (0.8) |
| age, mean (SD) | 61.8<br>(7.8) | 64.2 (7.4) | 62.1 (7.7) | 61.0 (7.8) | 59.9 (7.7) |
| **Education, n (%)** |  |  |  |  |  |
| *College or University degree* | 30237<br>(44.1) | 6681 (38.9) | 7267 (42.5) | 7848 (45.8) | 8441 (49.2) |
| *A levels/AS levels or*<br>*equivalent* | 9162<br>(13.4) | 2255 (13.1) | 2316 (13.5) | 2319 (13.5) | 2272 (13.3) |
| *O levels/GCSEs or*<br>*equivalent* | 14001<br>(20.4) | 3820 (22.2) | 3616 (21.1) | 3446 (20.1) | 3119 (18.2) |
| *CSEs or equivalent* | 2804<br>(4.1) | 673 (3.9) | 696 (4.1) | 688 (4.0) | 747 (4.4) |
| *NVQ or HND or HNC or*<br>*equivalent* | 3512<br>(5.1) | 957 (5.6) | 924 (5.4) | 842 (4.9) | 789 (4.6) |
| *Other professional*<br>*qualifications e.g nursing,*<br>*teaching* | 3327<br>(4.9) | 1018 (5.9) | 846 (4.9) | 792 (4.6) | 671 (3.9) |
| White ethnicity, n (%) | 66357<br>(96.8) | 16668<br>(97.0) | 16522<br>(96.6) | 16574<br>(96.7) | 16593<br>(96.8) |
| diet, mean (SD) <sup>b</sup> | 7.9 (4.4) | 7.8 (4.4) | 7.8 (4.3) | 7.9 (4.4) | 8.1 (4.6) |
| Discretionary screen time,<br>mean (SD) <sup>c</sup> | 3.6 (2.1) | 4.1 (2.3) | 3.7 (2.1) | 3.5 (2.1) | 3.2 (2.0) |
| **Medication, n (%)** |  |  |  |  |  |
| *Cholesterol, n (%)* | 7927<br>(11.6) | 2977 (17.3) | 2055 (12.0) | 1587 (9.3) | 1308 (7.6) |
| *Blood Pressure, n (%)* | 5792<br>(8.4) | 2075 (12.1) | 1498 (8.8) | 1234 (7.2) | 985 (5.7) |
| *Diabetes, n (%)* | 83 (0.1) | 21 (0.1) | 20 (0.1) | 21 (0.1) | 21 (0.1) |
| sleep (hours per day), (mean<br>(SD)) | 7.17<br>(0.98) | 7.19 (1.06) | 7.16 (0.98) | 7.16 (0.94) | 7.16 (0.92) |
| smokers, n (%) | 28659<br>(41.8) | 7744 (45.1) | 7170 (41.9) | 6964 (40.6) | 6781 (39.6) |
| Family history of CVD, n (%) | 37362<br>(54.5) | 9881 (57.5) | 9380 (54.9) | 9128 (53.3) | 8973 (52.4) |
| Family history of cancer n (%) | 21207<br>(30.9) | 5475 (31.9) | 5339 (31.2) | 5229 (30.5) | 5164 (30.1) |
| All-cause mortality, n (%) | 2134<br>(3.1) | 839 (4.9) | 505 (3.0) | 442 (2.6) | 348 (2.0) |
| CVD mortality, n (%) | 575<br>(0.8) | 265 (1.5) | 117 (0.7) | 110 (0.6) | 83 (0.5) |
| **Daily Durations across intensities and methods <sup>d</sup>** |  |  |  |  |  |
| *ML_MVPA (mean (SD))* | 38.3<br>(26.5) | 20.3 (13.9) | 31.8 (17.6) | 42.3 (22.3) | 58.7 (31.6) |
| *2LC_MVPA (mean (SD))* | 12.2<br>(14.5) | 2.3 (2.0) | 6.4 (4.1) | 12.6 (8.0) | 27.3 (19.8) |
| *1LC_MVPA (mean (SD))* | 36.0<br>(30.2) | 7.2 (3.5) | 19.6 (4.0) | 37.3 (6.7) | 79.9 (23.5) |
| *ML_MPA (mean (SD))* | 33.0<br>(24.0) | 18.1 (12.6) | 27.6 (16.1) | 36.1 (20.6) | 50.2 (30.0) |
| *2LC_MPA (mean (SD))* | 8.5 (9.4) | 1.8 (1.5) | 4.8 (3.0) | 9.0 (5.4) | 18.4 (12.4) |
| *1LC_MPA (mean (SD))* | 25.3<br>(19.9) | 5.7 (2.8) | 14.8 (3.1) | 26.9 (5.1) | 53.8 (15.3) |
| *ML_VPA (mean (SD))* | 5.3 (6.0) | 2.2 (3.3) | 4.2 (4.5) | 6.1 (5.7) | 8.6 (7.7) |
| *2LC_VPA (mean (SD))* | 3.7 (5.4) | 0.6 (0.6) | 1.7 (1.4) | 3.6 (3.0) | 8.9 (8.2) |
| *1LC_VPA (mean (SD))* | 10.7<br>(11.0) | 1.6 (1.0) | 4.8 (1.8) | 10.4 (3.4) | 26.2 (10.5) |
| *ML_LPA (mean (SD))* | 119.4<br>(62.9) | 108.9<br>(59.9) | 118.9<br>(62.5) | 123.4<br>(63.2) | 126.3 (64.6) |
| *2LC_LPA (mean (SD))* | 116.7<br>(63.4) | 76.2 (43.0) | 107.2<br>(51.6) | 128.8<br>(58.7) | 154.7 (69.2) |
| *1LC_LPA (mean (SD))* | 254.3<br>(79.8) | 199.8<br>(67.1) | 250.3<br>(68.4) | 274.9<br>(73.0) | 292.4 (78.2) |

<sup>a</sup> Quartiles were categorized from low to high based on the one-level cadence method derived daily MVPA duration. This table was based on the 95th percentile of the one-level cadence derived MVPA sample.
<sup>b</sup> fruit and vegetables servings per day
<sup>c</sup> time spent watching TV and computer per day
Abbreviations: CVD, cardiovascular diseases; MVPA, moderate-to-vigorous intensity physical activity; MPA, moderate intensity physical activity; VPA, vigorous intensity physical activity; LPA, light intensity physical activity; machine learning method (ML), one-level cadence method (1LC), two-level cadence method (2LC); SD, standard deviation;

**Supplementary table 2.**
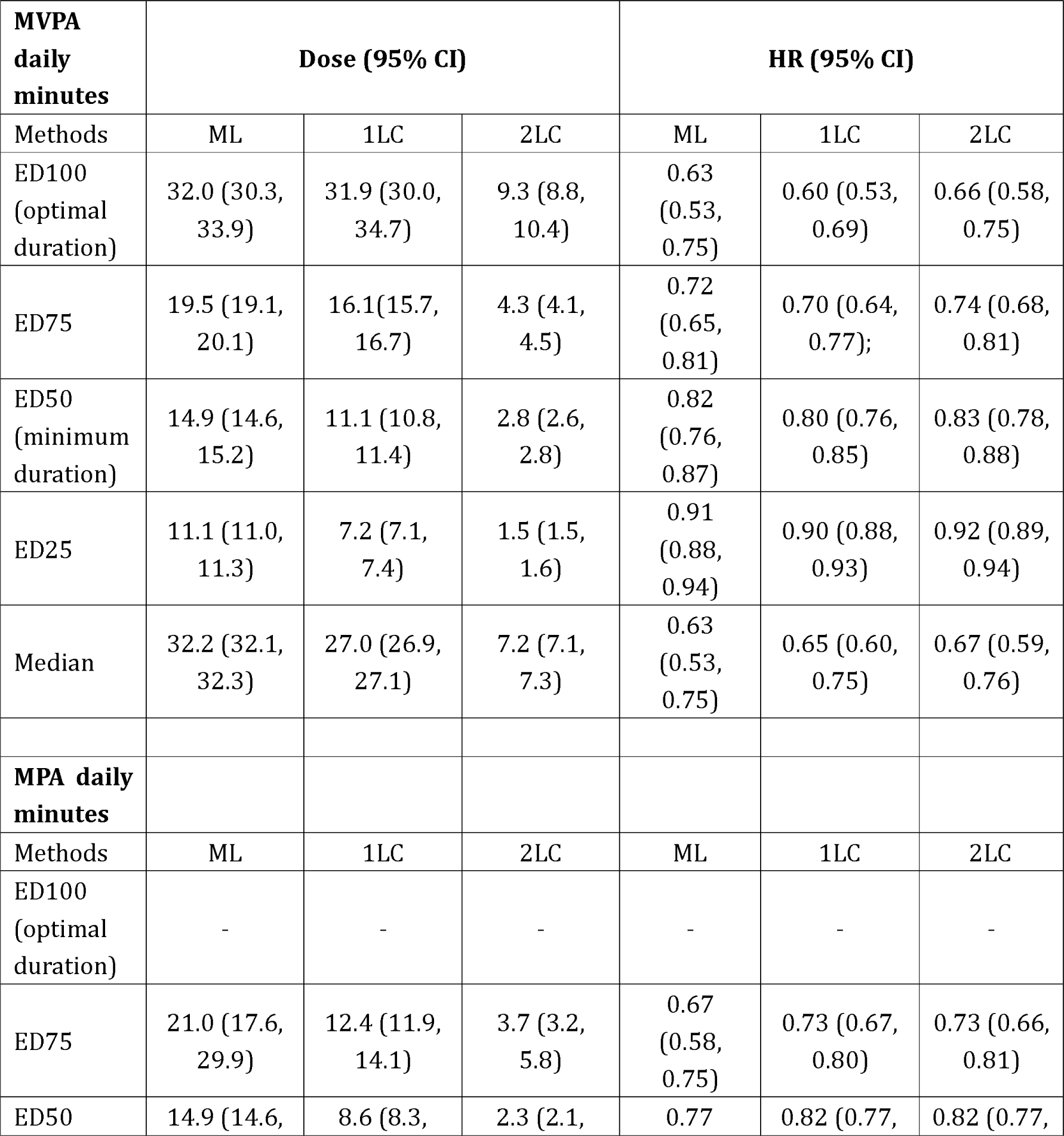
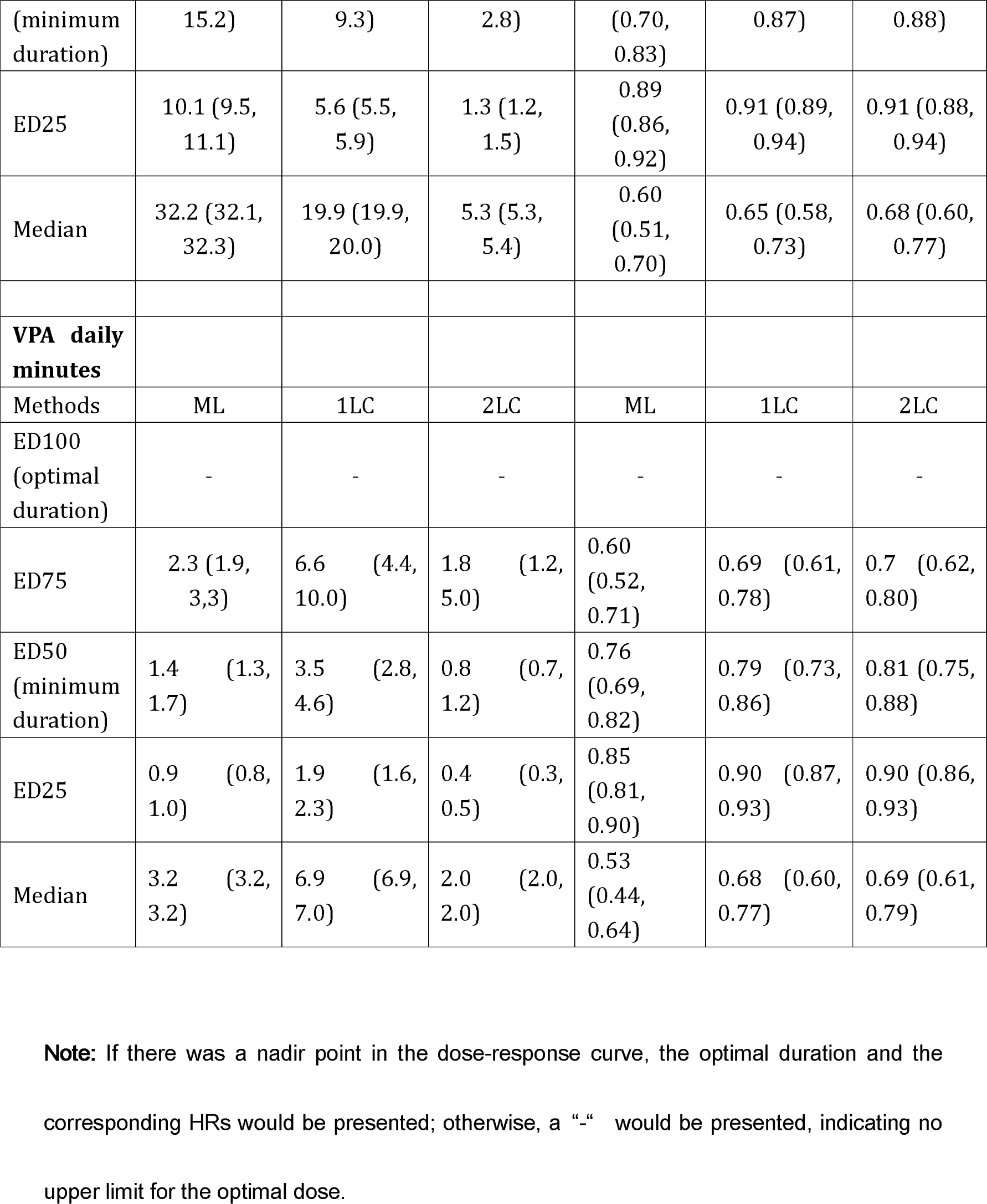
The duration associated with the ED25, ED50, ED75, ED100 and the median of the intensities duration derived from cadence-based methods (1LC and 2LC)* and machine learning (ML) method with all-cause mortality

**Supplementary Table 3.** The duration associated with the ED25, ED50, ED75, ED100 and the median of the intensities duration derived from cadence-based methods (1LC and 2LC)* and machine learning (ML) method with CVD mortality

| **MVPA daily minutes** | **Dose (95% CI)** |  |  | **HR (95% CI)** |  |  |
| --- | --- | --- | --- | --- | --- | --- |
| Methods | ML | 1LC | 2LC | ML | 1LC | 2LC |
| ED100 (optimal duration) | - | 32.0 (30.0, 34.7) | 18.0 (10.2, 19.9) | - | 0.61 (0.47, 0.78) | 0.58 (0.48, 0.76) |
| ED75 | 20.0 (19.0, 23.5) | 15.9 (15.1, 18.7) | 5.6 (4.1, 9.9) | 0.60 (0.50, 0.71) | 0.71 (0.59, 0.84) | 0.69 (0.58, 0.83) |
| ED50 (minimum duration) | 15.0 (14.5, 16.6) | 10.7 (10.3, 12.0) | 3.0 (2.5, 3.9) | 0.75 (0.68, 0.82) | 0.81 (0.72, 0.90) | 0.76 (0.66, 0.87) |
| ED25 | 11.1 (10.8, 11.8) | 7.0 (6.7, 7.5) | 1.6 (1.4, 1.9) | 0.86 (0.82, 0.91) | 0.90 (0.86, 0.95) | 0.89 (0.84, 0.94) |
| Median | 32.2 (32.1, 32.3) | 27.0 (26.9, 27.1) | 7.2 (7.1, 7.3) | 0.49 (0.40, 0.61) | 0.61 (0.48, 0.79) | 0.59 (0.46, 0.77) |
| **MPA daily minutes** |  |  |  |  |  |  |
| Methods | ML | 1LC | 2LC | ML | 1LC | 2LC |
| ED100 (optimal duration) | - | - | 6.4 (5.4, 8.2) | - | - | 0.64 (0.47, 0.87) |
| ED75 | 17.4 (16.8, 22.3) | 13.6 (11.4, 19.9) | 3.0 (2.8, 3.4) | 0.65 (0.55, 0.77) | 0.69 (0.56, 0.84) | 0.73 (0.60, 0.88) |
| ED50 (minimum duration) | 12.9 (12.5, 14.9) | 9.1 (8.0, 12.6) | 1.9 (1.8, 2.1) | 0.77 (0.69, 0.85) | 0.79 (0.70, 0.90) | 0.82 (0.73, 0.92) |
| ED25 | 9.4 (9.2, 10.2) | 5.5 (5.3, 7.0) | 1.1(1.1, 1.2) | 0.88 (0.84, 0.93) | 0.90 (0.84, 0.95) | 0.91 (0.87, 0.96) |
| Median | 27.2 (27.1, 27.3) | 19.9 (19.9, 20.0) | 5.3 (5.3, 5.4) | 0.54 (0.43, 0.68) | 0.59 (0.43, 0.81) | 0.64 (0.48, 0.86) |
| **VPA daily minutes** |  |  |  |  |  |  |
| Methods | ML | 1LC | 2LC | ML | 1LC | 2LC |
| ED100 (optimal duration) | - | 8.2 (7.1, 9.5) | - | - | 0.61 (0.44, 0.85) | - |
| ED75 | 13.1 (4.1, 13.4) | 3.9 (3.7, 4.2) | 1.2 (0.4, 2.1) | 0.55 (0.44, 0.69) | 0.71 (0.58, 0.86) | 0.60 (0.49, 0.75) |
| ED50 (minimum duration) | 9.0 (7.4, 9.3) | 2.5 (2.4, 2.7) | 0.6 (0.4, 0.9) | 0.70 (0.60, 0.83) | 0.81 (0.72, 0.91) | 0.75 (0.67, 0.85) |
| ED25 | 5.3 (3.8, 5.7) | 1.5 (1.4, 1.6) | 0.3 (0.1, 0.4) | 0.86 (0.80, 0.92) | 0.90 (0.85, 0.96) | 0.86 (0.81, 0.92) |
| Median | 3.2 (3.2, 3.2) | 6.9(6.9, 7.0) | 2 (1.9, 2.0) | 0.59 (0.47, 0.75) | 0.62 (0.45, 0.83) | 0.52 (0.40, 0.69) |

**Note:** If there was a nadir point in the dose-response curve, the optimal duration and the corresponding HRs would be presented; otherwise, a “-” would be presented, indicating no upper limit for the optimal dose.

**Supplementary table 4.**
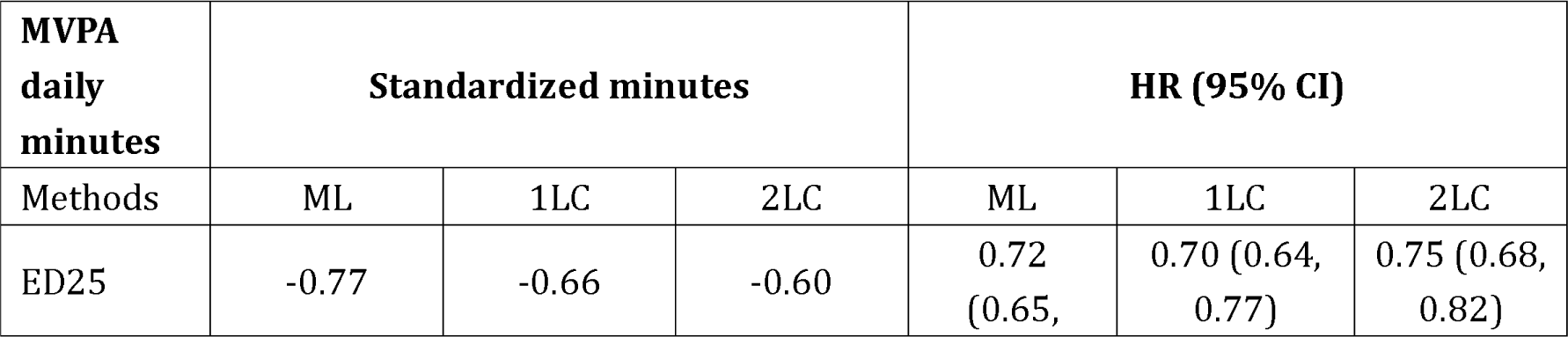
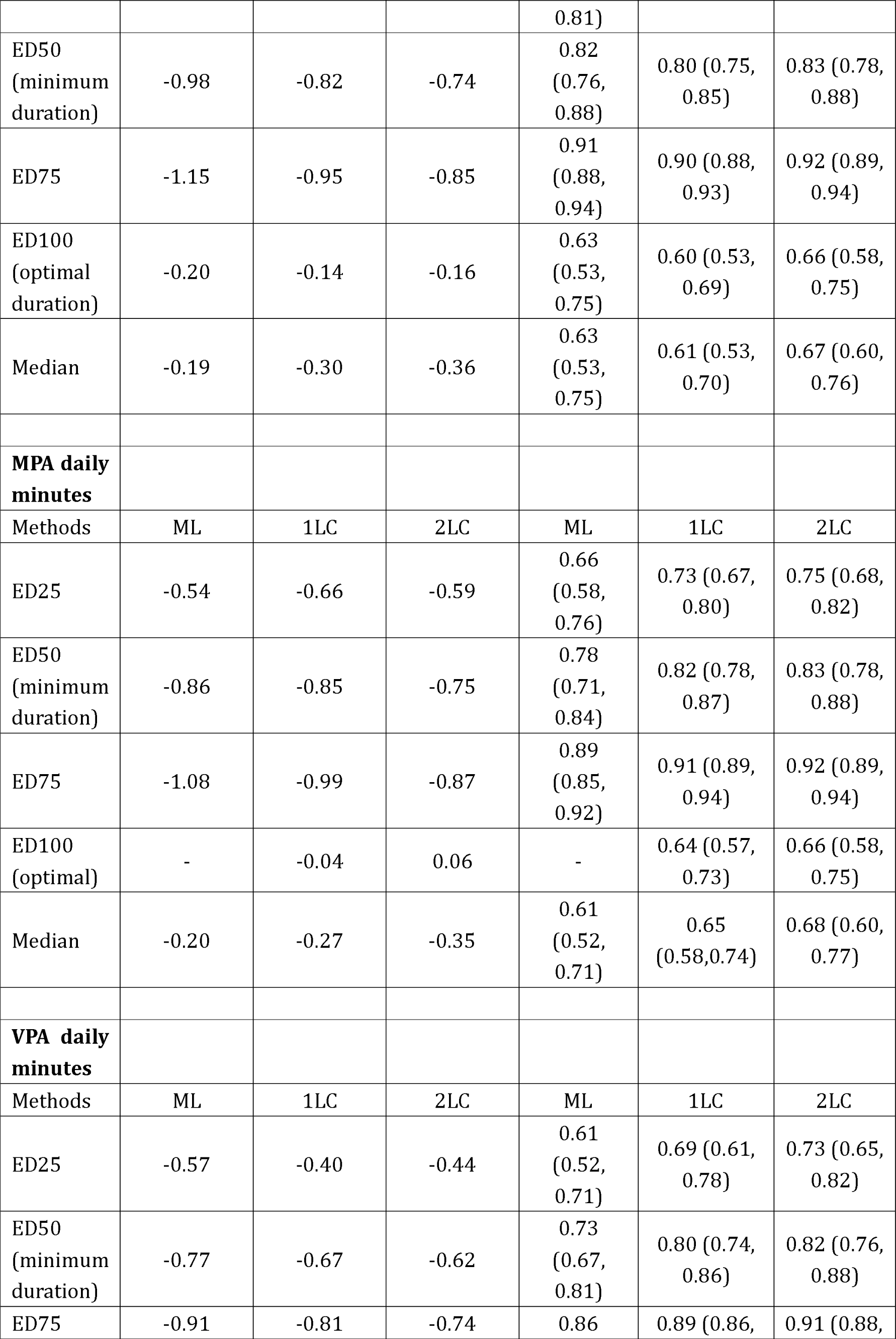
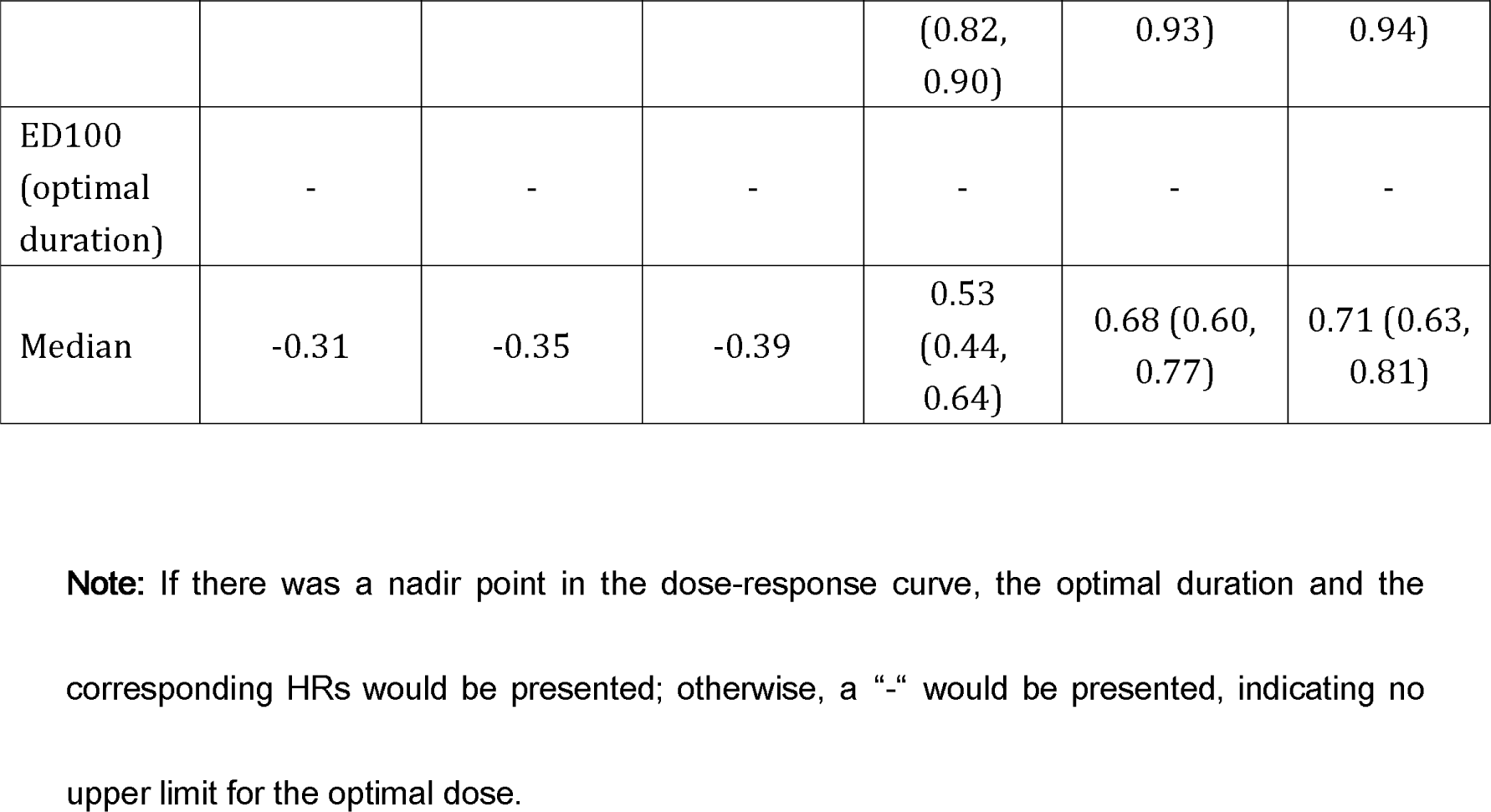
The standardized duration associated with ED25, ED50, ED75, ED100 and the median of the intensities duration derived from cadence-based methods (1LC and 2LC)* and machine learning (ML) method with all-cause mortality

**Supplementary table 5.**
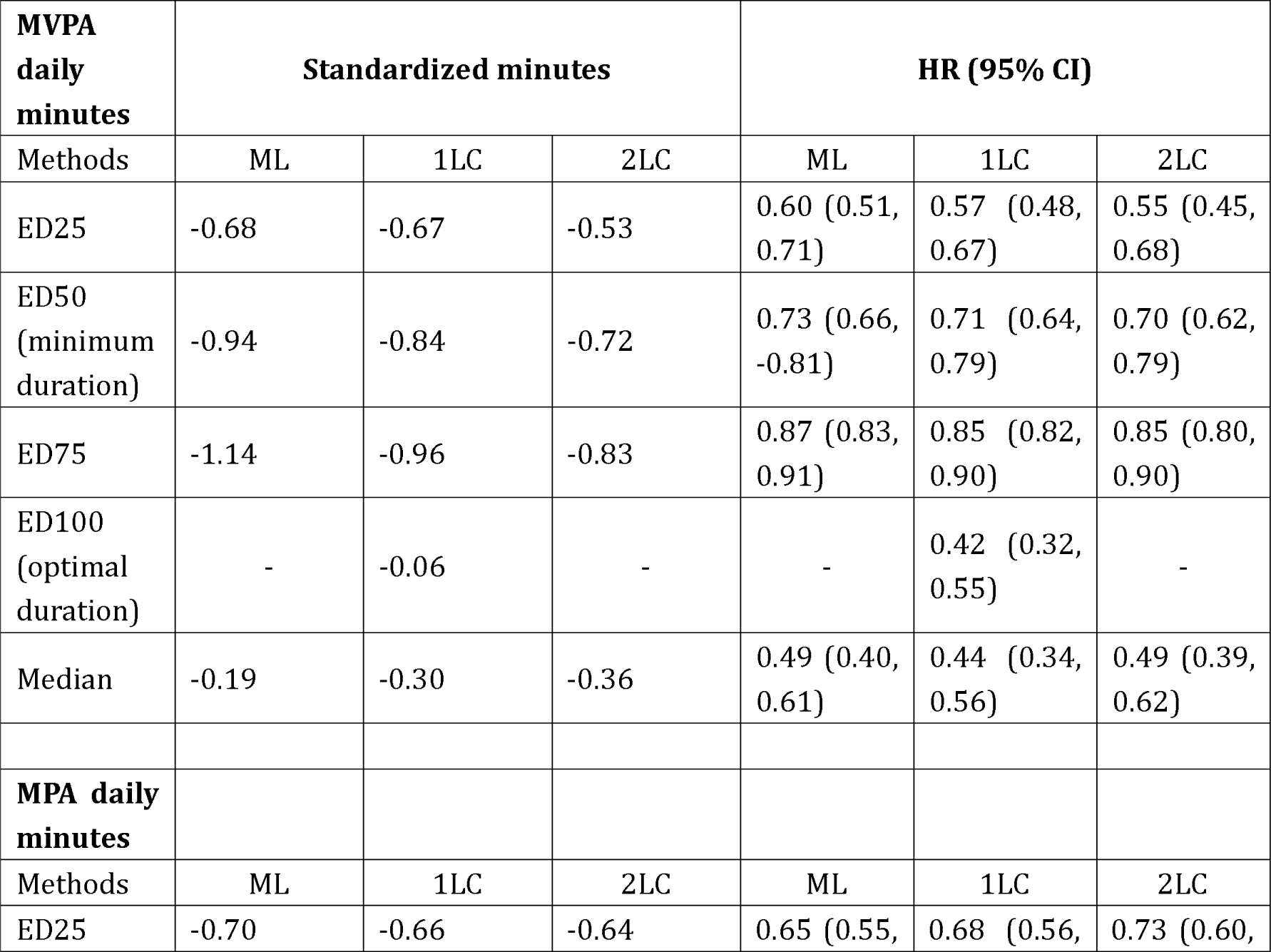
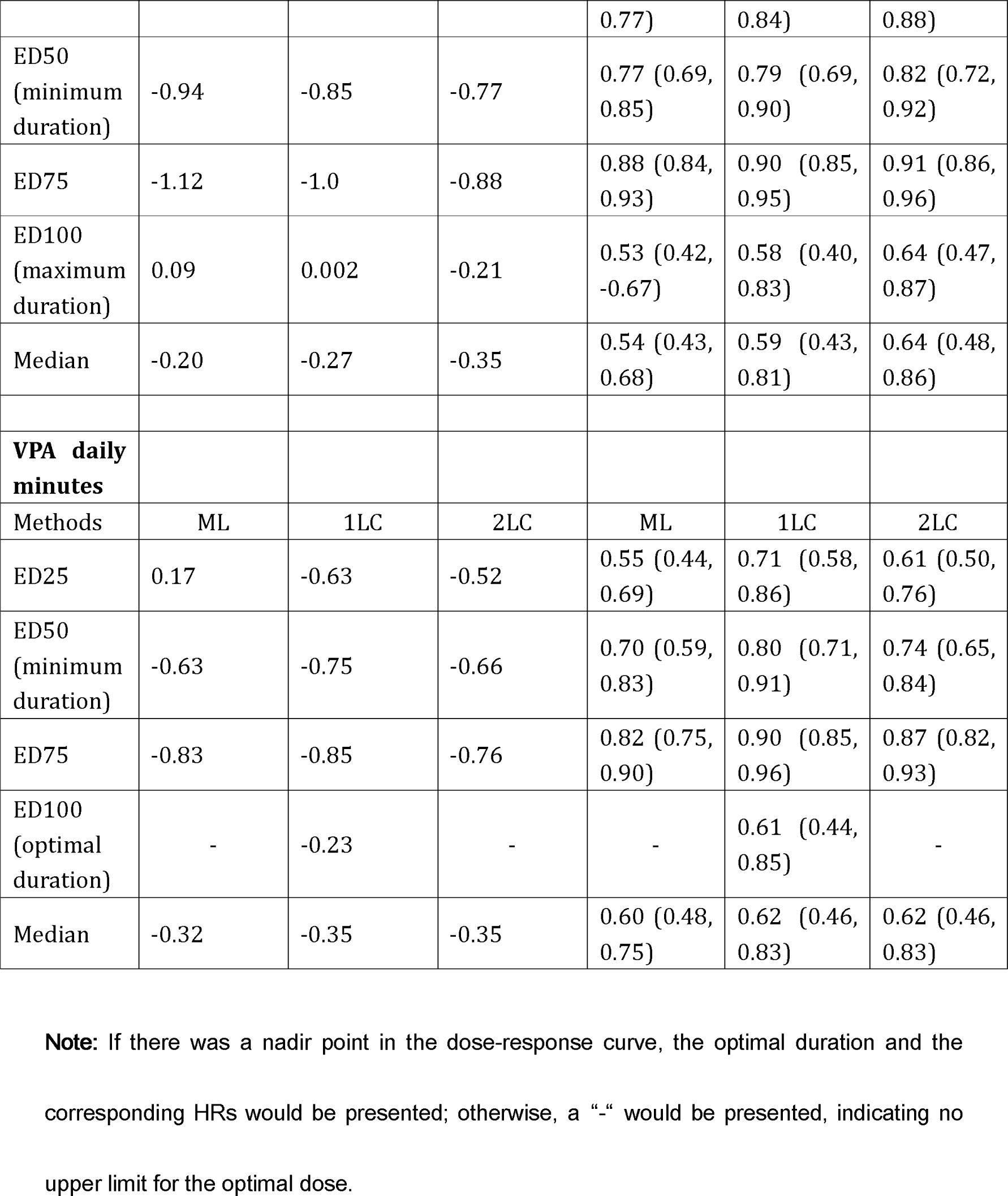
The standardized duration associated with the ED25, ED50, ED75, ED100 and the median of the intensities duration derived from cadence-based methods (1LC and 2LC)* and machine learning (ML) method with CVD mortality

